# External validation of ultrasound-based models for discrimination between benign and malignant adnexal masses in Italy: the prospective multicenter IOTA phase 6 study

**DOI:** 10.1101/2024.12.23.24319517

**Authors:** Francesca Moro, Marina Momi, Valentina Bertoldo, Ashleigh Ledger, Lasai Barreñada, Jolien Ceusters, Davide Sturla, Fabio Ghezzi, Elisa Mor, Letizia Fornari, Antonella Vimercati, Saverio Tateo, Marianna Roccio, Rosalba Giacchello, Roberta Granese, Daniela Garbin, Tiziana De Grandis, Federica Piccini, Patrizia Favaro, Olga Petruccelli, Anila Kardhashi, Ilaria Pezzani, Patrizia Ragno, Laura Falchi, Bruna Anna Virgilio, Erika Fruscella, Tiziana Tagliaferri, Annibale Mazzocco, Floriana Mascilini, Francesca Ciccarone, Federica Pozzati, Wouter Froyman, Ben Van Calster, Tom Bourne, Dirk Timmerman, Giovanni Scambia, Lil Valentin, Antonia Carla Testa

**Author notes:** **Corresponding authors:** Francesca Moro UniCamillus, International Medical University, Rome, Italy Fondazione Policlinico Universitario A. Gemelli IRCCS, Rome, Italy. The last two authors contributed equally to the work.

## Abstract

**Objective:** To prospectively validate the performance of the Risk of Malignancy Index (RMI), International Ovarian Tumor Analysis (IOTA) Simple Rules Risk Model (SRRisk), IOTA Assessment of Different NEoplasias in the adneXa (ADNEX) and the IOTA two-step strategy in different types of ultrasound centers in Italy.

**Methods:** This is a multicenter prospective observational study including regional referral centers and district hospitals in Italy. Consecutive patients with an adnexal mass examined with ultrasound by an IOTA certified ultrasound examiner with different levels of experience were included, provided they underwent surgery < 180 days after the inclusion scan. Ultrasound examination was performed transvaginally or transrectally and/or transabdominally based on the characteristics of the women and masses. Reference standard was the histology of the adnexal mass following surgical removal. Discrimination (area under receiver operating characteristic curve, AUROC), calibration, and clinical utility were assessed to illustrate the diagnostic performance of the methods. The performance of the models was also evaluated in predefined subgroups based on menopausal status, type of center (oncology vs non-oncology) and ultrasound examiner’s experience: [<500 scans performed, 500-5000 scans performed, >5000 scans performed; European Federation of Societies for Ultrasound in Medicine and Biology (EFSUMB) Level 1, Level 2, Level 3].

**Results:** 1567 patients were recruited between May 2017 and March 2020 from 23 italian centers. After data cleaning and application of exclusion criteria, our study population consisted of 1431 patients in 21 italian centers (10 oncological and 11 non-oncological). Based on histology, 995/1431 (69.5%) tumors were benign and 436/1431 (30.5%) were malignant (115/1431, 8.0% borderline, 263/1431, 18.4% primary invasive, 58/1431, 4.1% metastatic tumors). For all IOTA models (SRRisk, ADNEX with and without CA125, two step strategy with and without CA125), the AUROC was between 0.91 (95% CI 0.88-0.93) and 0.92 (0.89-0.94). The AUROC was 0.85 (0.81-0.87) for RMI. The malignancy risk was slightly underestimated by all IOTA models, but least so by SRRisk. All IOTA models had higher net benefit than RMI at risk thresholds from 1% to 50%. AUROC was ≥0.90 for all IOTA models in all subgroups, while it ranged from 0.84 to 0.90 for RMI.

**Conclusions:** SRRisk, ADNEX and the two step strategy with or without CA125 had similar and good ability to distinguish benign from malignant adnexal tumours in patients examined by either expert or non-expert ultrasound operators in Italy. Their discriminative performance and clinical utility was superior to that of RMI.

## Introduction

Ovarian cancer is the leading cause of death in women diagnosed with gynecological cancers.^1^ Most ovarian cancers are diagnosed at an advanced stage and require treatment in high volume centers by doctors with expertise in gynecological oncological surgery to optimize outcome.^2–4^ Correct preoperative characterization of adnexal masses is essential to decide on optimal management: clinical and ultrasound follow-up, surgery in a local center, or referral to an oncology center.^5,6^ Transvaginal ultrasound is the first line method for characterizing adnexal masses. If performed by an expert, subjective assessment of the ultrasound images is the optimal method for distinguishing benign from malignant masses.^7–9^ For less experienced ultrasound examiners there are other methods. The Risk of Malignancy Index (RMI) is a scoring system using clinical and ultrasound information that can be used to estimate the likelihood of an ovarian mass being malignant.^10^ In some European countries RMI is widely used to triage women with an adnexal mass for referral to an oncological center.^11–15^ The International Ovarian Tumor Analysis (IOTA) group has developed several ultrasound based methods that can be used to discriminate between benign and malignant adnexal masses: the Benign Descriptors,^16,17^, the Simple Rules, ^18^ four mathematical models to calculate the individual risk of malignancy in an adnexal mass (logistic regression model 1, LR1, logistic regression model 2, LR2, the Simple Rules Risk model, SRRisk, and Assessment of Different NEoplasias in the adneXa - ADNEX). ^19–21^ ADNEX is a multinomial regression model that calculates the probability of five outcome categories (benign, borderline, stage I primary invasive ovarian malignancy, stage II-IV primary invasive ovarian malignancy, and metastasis from another primary tumor). ^21^ ADNEX can be used with or without CA125 as predictor. ^21^ The IOTA group now recommends the IOTA two-step strategy, which means that first the Benign Descriptors are applied, and if these do not apply, ADNEX is used. ^17^

The diagnostic performance of the IOTA methods and of RMI has been validated in prospective and retrospective studies, but most validation studies tested the performance in the hands of experienced ultrasound examiners.^17,22–32^ No prospective study included examiners with little ultrasound experience and few included examiners with different levels of experience.^33–35^

The primary aim of this study is to prospectively validate the diagnostic performance of RMI, SRRisk, ADNEX and the IOTA two-step strategy in different types of ultrasound centers in Italy both overall and in relevant subgroups. The secondary aims are to explore the multinomial discrimination performance of ADNEX and the two step strategy, to validate the ability of the Benign Descriptors to correctly classify an adnexal mass as benign, and to validate the classification performance of the Simple Rules and of subjective assessment overall and in the subgroups based on level of ultrasound experience.

## Methods

### Study design and participants

This is an italian multicenter prospective external validation study of ultrasound based models to discriminate between benign and malignant adnexal masses. The protocol was approved by the Ethical Committee of the Fondazione Policlinico A. Gemelli, IRCCS (PROT 27665/16) and of each participating center (Appendix 1). Written informed consent was obtained from all patients. The study was conducted in accordance with the TRIPOD cluster guidelines.

### Patients

Consecutive patients with a known or suspected adnexal mass examined with ultrasound by an IOTA certified ultrasound examiner^36^ and confirmed to have an adnexal mass judged not to be physiological were eligible for inclusion provided they were expected to undergo surgical removal of the mass. The patients were collected between May 2017 and March 2020. Exclusion criteria were: patient’s age <18 years, pregnant patients, patients with previous bilateral adnexectomy, patients examined in centers that recruited < 10 patients, only transabdominal ultrasound performed, surgery performed more than 180 days after the ultrasound examination, and denial or withdrawal of informed consent.

### Data collection

Information on age, parity, menopausal status and indication for the ultrasound examination was prospectively collected, as well as information on type of hospital (private practice, local public hospital, regional public hospital, or university hospital), type of center (oncological vs non-oncological), and type of ultrasound center (general gynecologic outpatient clinic or specialized ultrasound center). An oncological center was defined as a tertiary referral center with a dedicated gynecological oncology unit. Information on the ultrasound system used, ultrasound examiner’s name and level of experience was also recorded. The level of ultrasound experience was based on the number of gynecological scans in non-pregnant women that the examiner had performed at the start of the study. Low experience was defined as <500 scans, intermediate experience as 500-5000 scans, and high experience as >5000 scans. We also recorded the level of experience according to the European Federation of Societies for Ultrasound in Medicine and Biology (EFSUMB level 1, 2 or 3 ^37^), and the number of ovarian masses that the operator had examined with ultrasound during the preceeding year (classified as <50, i.e. < 1 per week; 50-200, i.e. up to 4 per week; >200, i.e. > 4 per week).

### Ultrasound examination

A standardized transvaginal (or transrectal if vaginal was not possible) ultrasound examination including color or power Doppler ultrasound examination was performed, supplemented with transabdominal ultrasound if transvaginal or transrectal ultrasound examination was not sufficient. The IOTA examination and measurement technique were used, and the ultrasound findings were described using the IOTA terminology.^38^ Information on all the variables required for the IOTA Benign Descriptors, Simple Rules, SRRisk, ADNEX and RMI were prospectively collected and recorded. Results of subjective assessment were recorded as benign, borderline or malignant. The degree of diagnostic confidence (certainly benign, probably benign, uncertain, probably malignant or probably borderline, certainly malignant or certainly borderline) as well as the specific diagnosis suggested by the ultrasound examiner and chosen from a list of pre-defined diagnoses were also recorded.

If more than one adnexal mass was present, only the one with the most complex ultrasound morphology was included in our statistical analysis. If the ultrasound morphology was similar in all masses, the largest one or the one most easily accessible with ultrasound was used in our statistical calculations. The management was decided by the referring clinician, who took into account clinical symptoms, ultrasound results based on subjective evaluation of the ultrasound images (i.e. those reported in the clinical ultrasound report), and results of other imaging modalities (e.g. computer tomography or magnetic resonance imaging), tumor markers, and patient’s preference.

### Reference standard

Reference standard was the histology of the adnexal mass following surgical removal within 180 days after the ultrasound examination by laparotomy or laparoscopy as considered appropriate by the surgeon. Borderline tumors were classified as malignant. The histology of the surgically removed tumor was determined at each local center. Central pathology review was not performed, because we found little differences between local and central pathology reports in a previous IOTA study.^19^ Pathologists were blinded to ultrasound predictor variables and model predictions but might have received information on the subjective assessment by the ultrasound examiner when clinically relevant. The stage of malignant tumors was recorded using the classification of the International Federation of Gynecology and Obstetrics (FIGO). ^39^

### Data cleaning

Data collection was done through the web-based clinical data miner (CDM) software.^40^ Patients automatically received a unique identifier upon enrolment. We encrypted all data communication to ensure data security. A team of statisticians and ultrasound examiners performed data cleaning. Data cleaning included sending queries to participating centers to retrieve missing information or to correct inconsistencies.

### Prediction models

We assessed the diagnostic performance of subjective assessment, RMI, Benign Descriptors, Simple rules, SRRisk, ADNEX with and without CA125, and the two-step strategy with and without CA125. Predictions were based on information obtained at the inclusion scan and so are blinded to the outcome. The results of the models were calculated after closing the study and were not used to guide patient managament.

*RMI* does not give an estimated risk but a non-negative integer (0 or higher), with higher scores suggesting a higher likelihood of malignancy. It includes clinical (CA125 and menopausal status) and ultrasound variables (multilocular cyst, solid areas, bilateral lesions, ascites, abdominal metastases).^10^

There are *four Benign Descriptors,* which classify tumors as benign: 1) a unilocular cyst with ground glass echogenicity and largest diameter <10 cm in a premenopausal woman is suggestive of endometrioma (BD 1); 2) a unilocular cyst with mixed echogenicity, acoustic shadows and largest diameter <10 cm in a premenopausal woman is suggestive of benign teratoma (BD 2); 3) a unilocular cyst with anechoic cyst fluid, smooth internal walls and largest diameter <10 cm in a pre- or post-menopausal woman is suggestive of a simple cyst or cystadenoma (BD3); 4) all other unilocular cysts with smooth internal walls and largest diameter <10 cm in a pre- or post-menopausal woman (BD 4) are suggestive of a benign cyst. ^17^

*Simple Rules* classify tumors as benign, inconclusive, or malignant based on the presence of five benign ultrasound features (unilocular cyst, smooth multilocular cyst with largest diameter <100 mm, acoustic shadows, solid component(s) are present but largest diameter is <7 mm, no vascularization on color Doppler) and five malignant ultrasound features (irregular solid tumor, irregular multilocular solid tumor with largest diameter ≥100 mm, at least 4 papillary projections, presence of ascites, very strong vascularization on color Doppler). The classification is inconclusive if none of the ten features is present, or if both benign and malignant features are present. ^18^ We added inconclusive tumors to those predicted to be malignant, resulting in a binary classifier (benign or malignant).

*SRRisk* is a logistic regression model that calculates the risk that an adnexal tumor is malignant based on type of center (oncology center vs other center) and on the ten binary ultrasound features used in the Simple Rules. ^20^

*ADNEX* is a multinomial regression model that calculates the probability of five outcome categories: benign, borderline, stage I primary invasive ovarian malignancy, stage II-IV primary invasive ovarian malignancy, and metastasis from another primary tumor. ^21^ One minus the probability of a benign tumor equals the estimated risk of malignancy. ADNEX includes three clinical variables (age, type of center, i.e. oncology vs non-oncology center, serum level of CA125) and six ultrasound variables (maximum diameter of the lesion in mm, proportion of solid tissue calculated as the maximum diameter of the largest solid component in mm divided by the maximum diameter of the lesion in mm, presence of more than 10 cyst locules, number of papillary projections, acoustic shadows, ascites). The variable CA125 is optional, but for multinomial discrimination ADNEX works better with than without CA125. ^22^

The *two-step strategy* uses the Benign Descriptors as a first step. If a Benign Descriptor applies, the mass is classified as benign, if not, ADNEX with or without CA125 is used to estimate the risk of malignancy. ^17^ Information about how we estimated the risk of malignancy when a Benign Descriptor applied is found in Appendix 2.

### Statistical analysis and sample size

We followed a prespecified statistical analysis plan. The statistical analyses were performed with R version 4.1.2. The adequacy of the sample size is discussed in Appendix 3. We report all performance measures with 95% confidence intervals (CI).

Despite it being strongly recommended to collect blood samples for measurement of serum CA125 in all patients, CA125 results were missing in some patients. Missing CA125 values were imputed. We performed multiple imputation using the method of fully conditional specification, generating 100 imputations, leading to 100 completed datasets. To estimate the missing values of CA125, we used predictive mean matching regression using the outcome and variables that are probably related to either the level of CA125 itself, or to the unavailability of CA125. The multiple imputation procedure is described in Appendix 4. To calculate the predictions for each model, we used the formula present in the original paper.

Results are presented as absolute frequency (percentage) for nominal variables and as median, interquartile range (IQR) and range (min-max) for continuous variables as appropriate. We report the percentage of tumors to which a Benign Descriptor applied and the outcome of masses to which a Benign Descriptor applied (pooled analysis).

We calculated center-specific area under the receiver operating characteristic curve (AUROC) to estimate the ability to discriminate between benign and malignant adnexal masses for RMI and the risk models (SRRisk, ADNEX, IOTA two-step strategy) and used meta-analysis to obtain the overall AUROC per model. The heterogeneity between centers was assessed by calculating 95% prediction intervals (PI). The meta-analysis procedure is described in Appendix 5.

We assessed calibration of the risk models by calculating observed over expected ratio (O:E). O:E is the ratio of the observed risk of having the outcome divided by the risk estimated by the model. An O:E higher than 1 indicates that the model underestimates the risk of malignancy, and an O:E lower than 1 indicates that the model overestimates the risk of malignancy. The ideal value is 1. ^41,42^ We also constructed flexible calibration curves using loess. ^43^ We calculated center-specific O:E and calibration curves and combined them using meta-analysis. For the meta-analysis of calibration curves, we combined 13 centers with small sample size or low prevalence of malignancy into four groups to avoid computational problems. The four groups were: Santorso, Foggia, Treviso (group 1); Messina, Carpi, Montebelluna (group 2); Verona, Firenze, Padova, Rome (group 3); and Bari B, Asti, Bolzano (group 4). The meta-analysis procedure is described in Appendix 5.

Clinical utility to decide which patients to refer for specialized oncological care was estimated using decision curve analysis for risk thresholds between 1% and 50%.^44^ Net benefit is a measure of clinical utility. To know if a model is clinically useful, we compare it to treat all and treat none (in this case to refer all or to refer none to an oncology center). A model is clinically useful if it is superior to both treat all and treat none. Because RMI does not provide risk estimates, for RMI we computed clinical utility at the following fixed RMI scores: 200 (a threshold often used clinically and recommened in several national guidelines^11–15^), 250, 100 and 25. We show overall decision curves calculated using meta-analysis of center-specific decision curves. The meta-analysis procedure is described in Appendix 5.

We calculated sensitivity, specificity, positive predictive value (PPV) and negative predictive value (NPV) for subjective assessment and the Simple Rules (inconclusive cases classified as malignant) and for the risk models at risk of malignancy cut-offs of 1%, 3%, 5%, 10%, 15%, 20%, 25%, 30%, 40%, and 50%. For RMI, we report classification performance for cut-offs 25, 100, 200, and 250. We calculated center-specific sensitivity and specificity and combined them using meta-analysis.^45^ Centers with no true positive (TP) and no false positive (FP) test results at a specific threshold were excluded from the meta-analysis of PPV for that threshold. Centers with no true negative (TN) and no false negative (FN) at a specific threshold were excluded from the meta-analysis of NPV for that threshold.

To estimate the multinomial performance of ADNEX and the two-step strategy, we computed the Polytomous Discrimination Index (PDI) and calculated the AUROC for each pair of outcome categories using the conditional risk method.^46–47^ To evaluate calibration, we computed O:E ratios per category. Due to the small numbers in most centers, we anticipated computational problems when attempting to perform meta-analysis for multinomial performance. Therefore, we used the pooled dataset to estimate the multinomial performance of ADNEX and the two-step strategy.

### Subgroup analyses

We calculated AUROC and O:E ratio for prespecified subgroups based on menopausal status, type of center (oncology vs non-oncology), and ultrasound examiner’s experience (<500 scans performed, 500-5000 scans performed, >5000 scans performed; EFSUMB Level 1, Level 2, and Level 3) using pooled data due to the small numbers in most centers.

## Results

A total of 1567 patients were recruited from 23 italian centers. After data cleaning and application of exclusion criteria, our study population consisted of 1431 patients in 21 italian centers (10 oncological and 11 non-oncological centers) (Figure 1 and Supplementary Table S1). Based on histology, 995/1431 (69.5%) tumors were benign and 436/1431 (30.5%) were malignant (115/1431, 8.0% borderline, 263/1431, 18.4% primary invasive, 58/1431, 4.1% metastatic tumors). Tumor outcome according to center is shown in Supplementary Table S1. Clinical, ultrasound and histological characteristics of the study population are summarized in Table 1. Specific histological diagnoses are shown in Supplementary Table S2. The median age of the patients was 52 years (IQR 40-62, range 18 to 88), and 745 patients (52%) were postmenopausal. The median of the maximum diameter of the lesion was 69 (IQR 48-100, range 9 - 400) mm, 281 (20%) patients had bilateral masses, and 120 (8%) patients had ascites. CA125 was missing in 28% (394/1431) of patients. The characteristics of our study population and those of the studies in which the RMI and IOTA models were developed are shown in Supplementary Table S3. Patients in our study population were more frequently postmenopausal than those in the development sets of the IOTA models, the Ca125 values (when available) were lower, while acoustic shadowing and absent color Doppler signals were more common. Multilocular cysts were less common than in the development data set of the Simple Rules, and Ca125 values were higher than in the development data set of RMI. The distribution of tumor outcome (i.e., prevalence of borderline, stage I primary invasive, stage II-IV primary invasive, metastatic tumors) in our study population is reasonably similar to that in the studies in which the IOTA models were developed.

**Figure 1.**
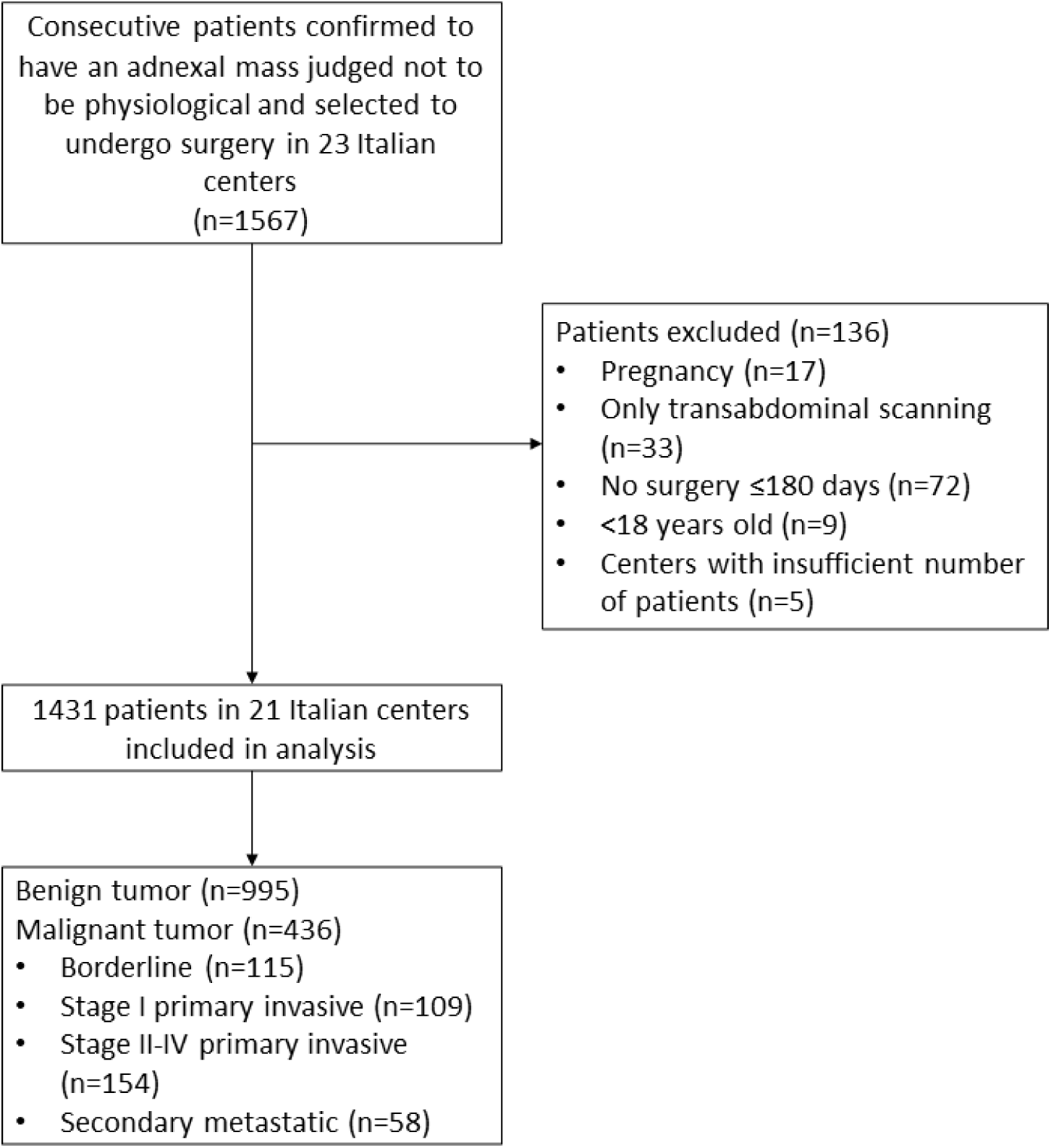
Flowchart of the patients included for the analysis.

**Table 1.**
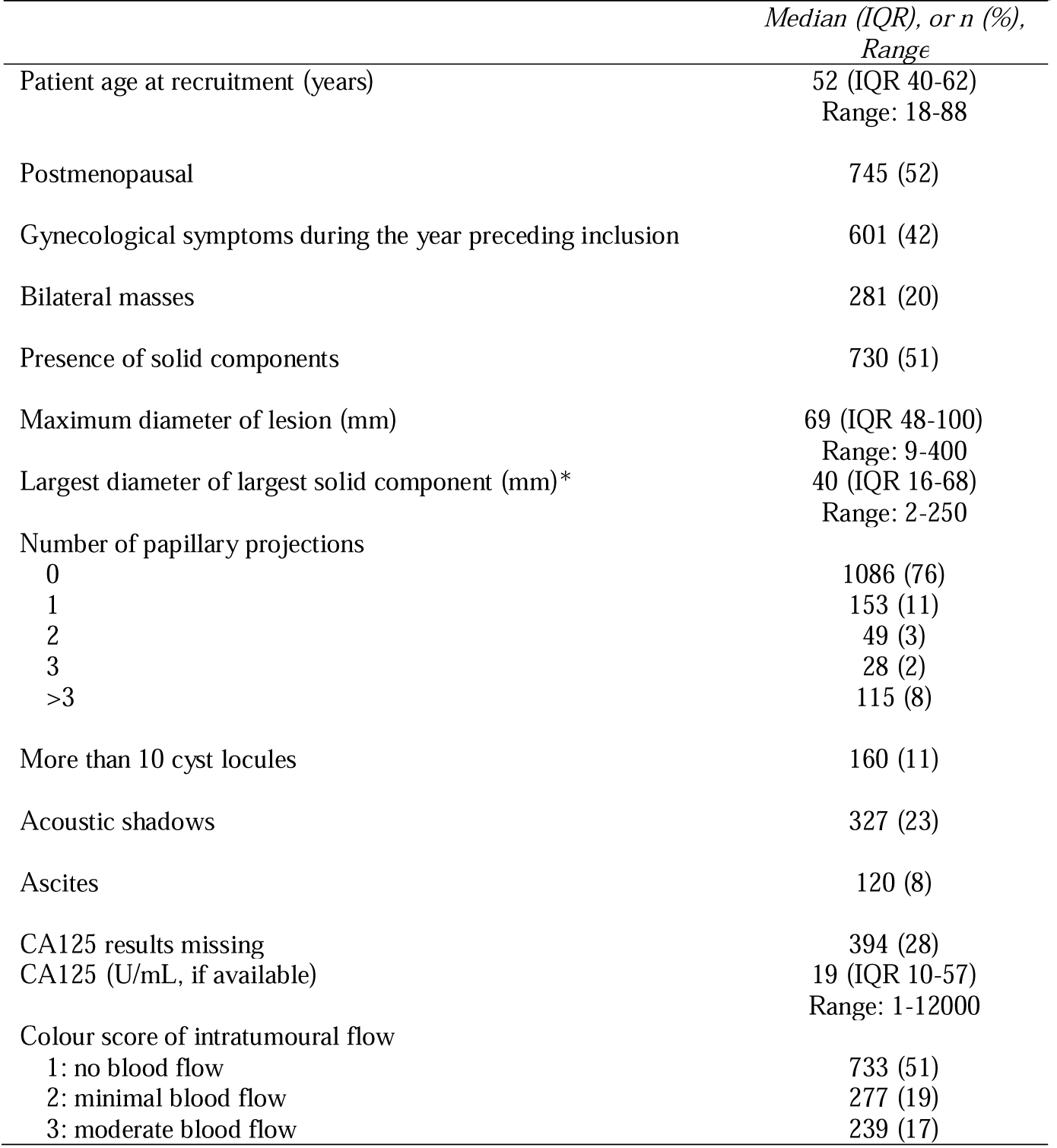
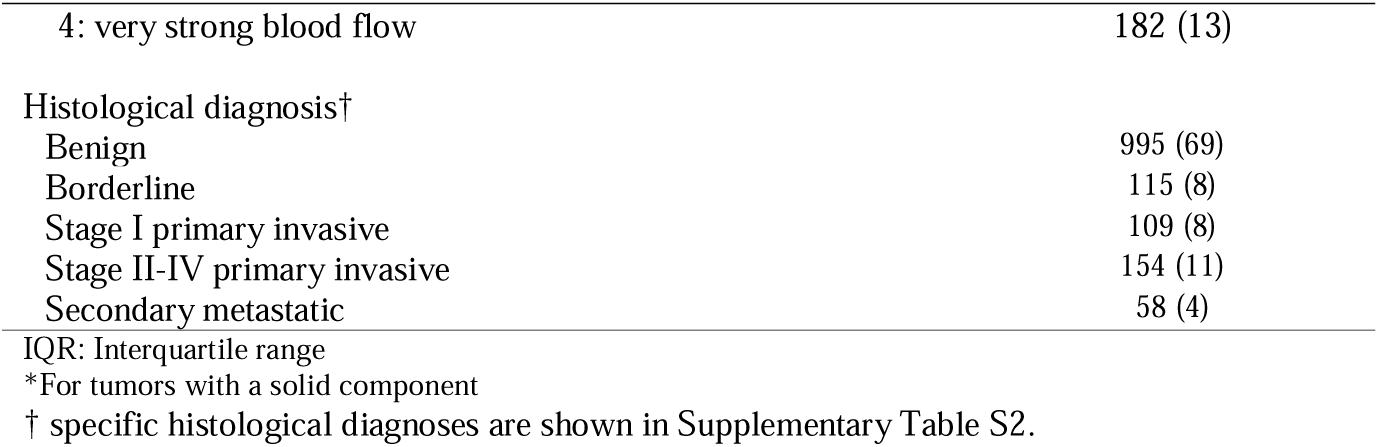
Clinical, ultrasound, and histological characteristics of the study population (n=1431).

|  | *Median (IQR), or n (%),*<br>*Range* |
| --- | --- |
| Patient age at recruitment (years) | 52 (IQR 40-62)<br>Range: 18-88 |
| Postmenopausal | 745 (52) |
| Gynecological symptoms during the year preceding inclusion | 601 (42) |
| Bilateral masses | 281 (20) |
| Presence of solid components | 730 (51) |
| Maximum diameter of lesion (mm) | 69 (IQR 48-100)<br>Range: 9-400 |
| Largest diameter of largest solid component (mm)* | 40 (IQR 16-68)<br>Range: 2-250 |
| Number of papillary projections |  |
| 0 | 1086 (76) |
| 1 | 153 (11) |
| 2 | 49 (3) |
| 3 | 28 (2) |
| >3 | 115 (8) |
| More than 10 cyst locules | 160 (11) |
| Acoustic shadows | 327 (23) |
| Ascites | 120 (8) |
| CA125 results missing | 394 (28) |
| CA125 (U/mL, if available) | 19 (IQR 10-57)<br>Range: 1-12000 |
| Colour score of intratumoural flow |  |
| 1: no blood flow | 733 (51) |
| 2: minimal blood flow | 277 (19) |
| 3: moderate blood flow | 239 (17) |
| 4: very strong blood flow | 182 (13) |
| Histological diagnosis† |  |
| Benign | 995 (69) |
| Borderline | 115 (8) |
| Stage I primary invasive | 109 (8) |
| Stage II-IV primary invasive | 154 (11) |
| Secondary metastatic | 58 (4) |

IQR: Interquartile range
\*For tumors with a solid component
† specific histological diagnoses are shown in Supplementary Table S2.

The BDs applied to 328/1431 (23%) tumors, of which 325 (99%) were benign, 3/328 (1%, 95% CI 1-2) were borderline, and none was an invasive malignany (Supplementary Table S4).

For all IOTA models (ADNEX with and without CA125, SSRisk, two step strategy with and without CA125), the overall AUROC was ≥0.91, while that of RMI was 0.85 (Figure 2). Differences in AUROC between centers (heterogeneity) were smallest for SRRisk (Figure 2, Supplementary Figure S1-S6).

**Figure 2.**
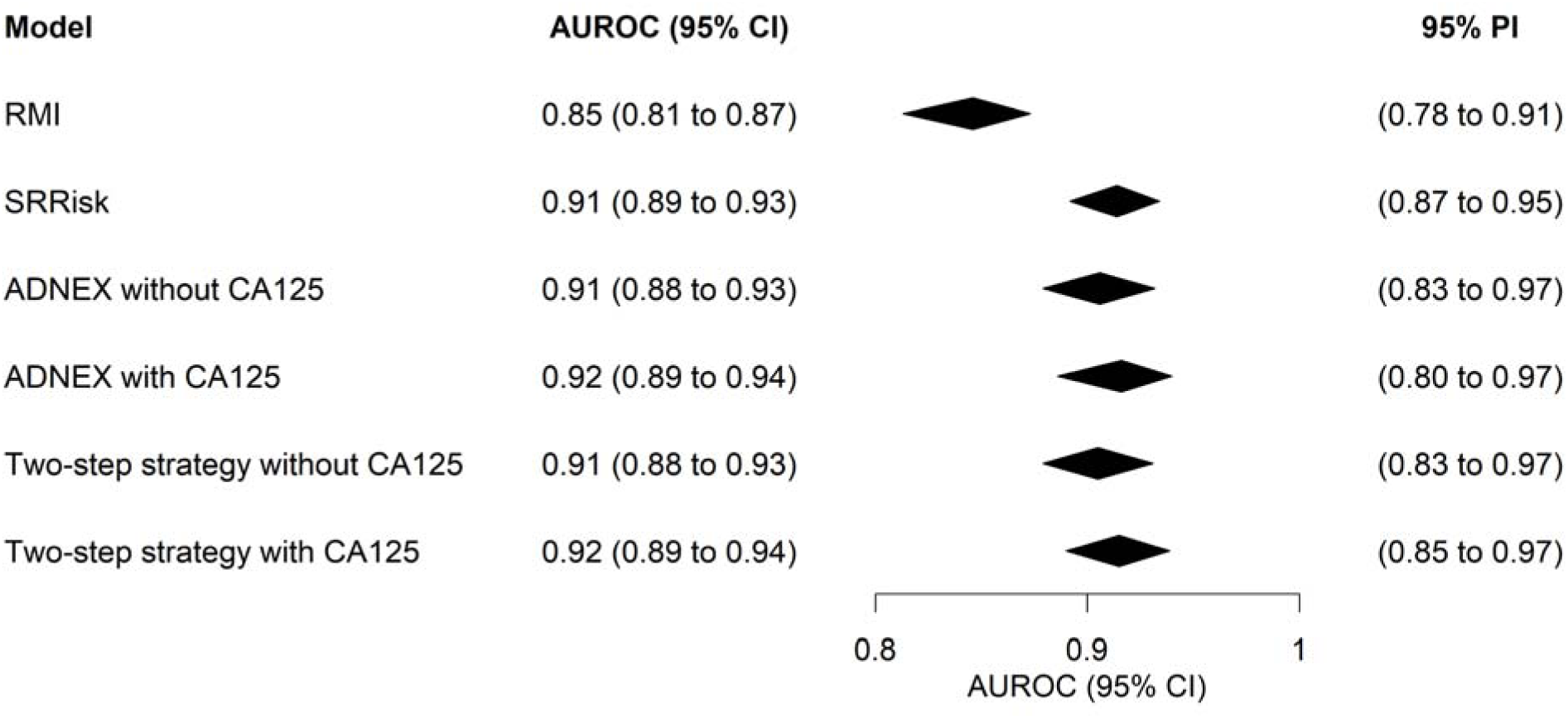
Summary forest plot of area under the receiver operating characteristic curve (AUROC) based on meta-analysis of data from 21 centers. CI; Confidence Interval, PI; Prediction Interval.

At a risk threshold of 10% (the risk threshold recommended in an international consensus statement for referring patients to an oncology center ^6^), all IOTA models (SRRisk, ADNEX with and without Ca125 and the two-step strategy with and without Ca125) had sensitivity >0.9 with specificity ranging from 0.77 to 0.80. ADNEX and the two step strategy had the same classification performance at risk threshold 10%: sensitivity 0.92 and specificity 0.80 when CA125 was included as a predictor, sensitivity 0.94 and specificity 0.77 when CA125 was not included as a predictor (Supplementary Table S5). At a threshold of 200, RMI had sensitivity 0.58 and specificity 0.94 (Supplementary Table S6). The Simple Rules were applicable in 1244/1431 (87%) tumors and had sensitivity 0.90 and specificity 0.85 (inconclusive cases classified as malignant). Subjective assessment had sensitivity 0.93 and specificity 0.88 (Supplementary Table S7).

The malignancy risk was slightly underestimated by all IOTA models (O:E ratios 1.04 −1.20) (Table 2). SRRisk was slightly better calibrated than other IOTA models (point estimate O:E ratio 1.04; 95% CI 0.97-1.12) (Table 2, Figure 3). The relation between the RMI score and the observed prevalence of malignancy is shown in supplementary Figure S7. At RMI score 200, the observed prevalence of malignancy was 55% (95% CI 49-61).

**Figure 3.**
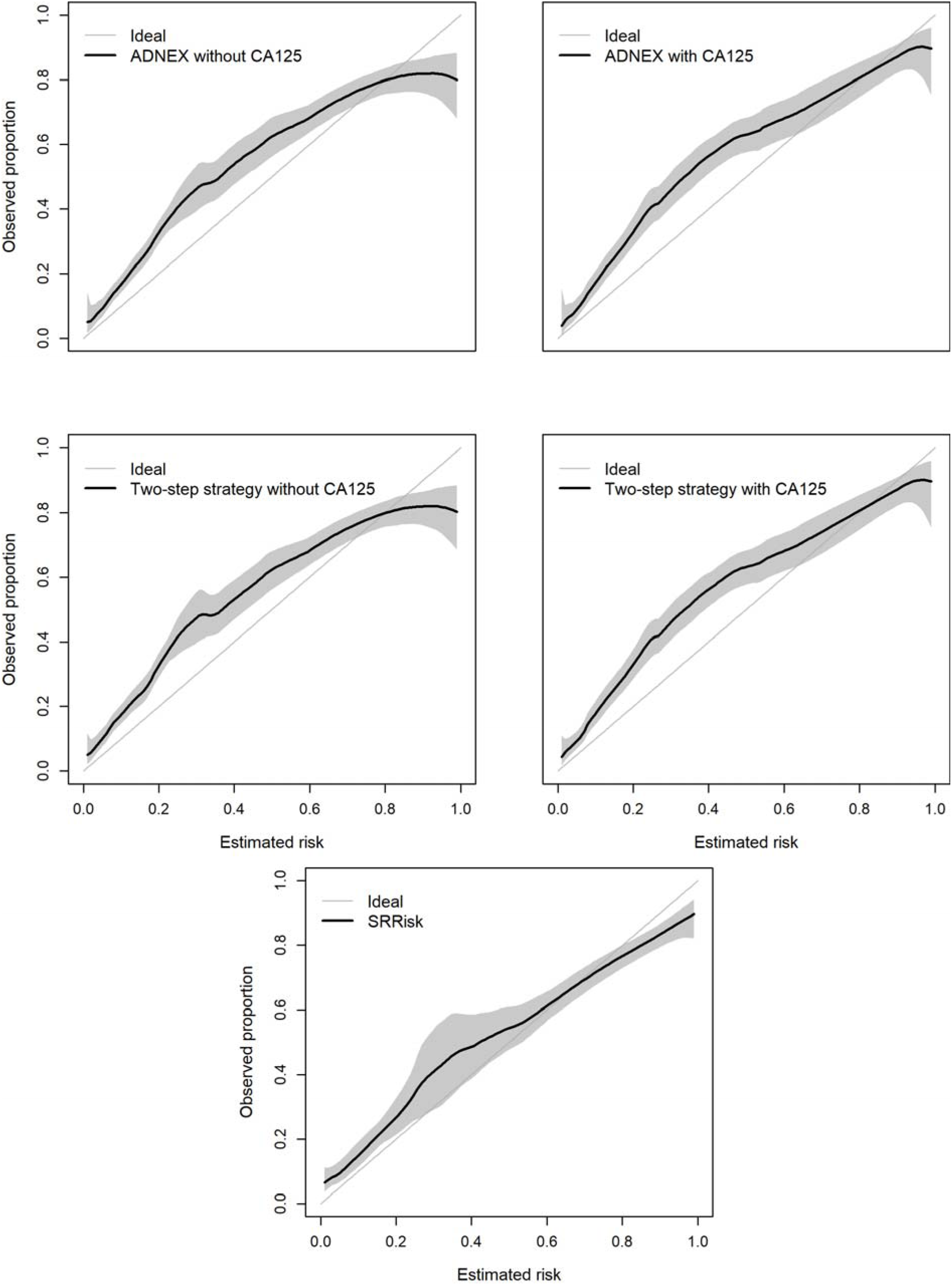
Flexible calibration curves using loess based on meta-analysis. Due to computational problems, we divided 13 centers with low sample size or low prevalence of malignancy into four groups: Santorso, Foggia, Treviso (group 1); Messina, Carpi, Montebelluna (group 2); Verona, Firenze, Padova, Roma (group 3); Bari B, Asti, Bolzano (group 4).The curves were obtained with meta-analysis of center-specific curves from eight centers and from the four groups. SRRisk; Simple Rules Risk Model, ADNEX; Assessment of Different NEoplasias in the adnexa.

**Table 2.** Calibration in terms of observed over expected ratio based on meta-analysis of data from.

| Model | O:E ratio (95% CI) |
| --- | --- |
| SRRisk | 1.04 (0.97; 1.12) |
| ADNEX without CA125 | 1.11 (1.04; 1.20) |
| ADNEX with CA125 | 1.18 (1.07; 1.29) |
| Two-step without CA125 | 1.13 (1.05; 1.21) |
| Two-step with CA125 | 1.20 (1.09; 1.32) |

O:E ratio, observed over expected ratio. Measure of calibration in the large (mean calibration) is calculated as the observed risk of having the outcome event in the entire validation dataset divided by the average risk predicted by the model. A value >1 indicates that the model is underestimating the average risk. A value <1 means that the model is over-estimating the average risk.
CI; Confidence Interval, SRRisk; Simple Rules Risk Model, ADNEX; Assessment of Different NEoplasias in the adnexa.

Decision curves are shown in Figure 4. Irrespective of which RMI cutoff was used, the IOTA methods had higher net benefit than RMI. At risk thresholds lower than 15%, ADNEX and the two-step strategy had the highest net benefit, at risk thresholds above 20% SRRisk and subjective assessment had highest net benefit. RMI at cutoff 200 was less clinically useful than simply treating everyone (referring everyone to an oncology center) at risk thresholds below 15%.

**Figure 4.**
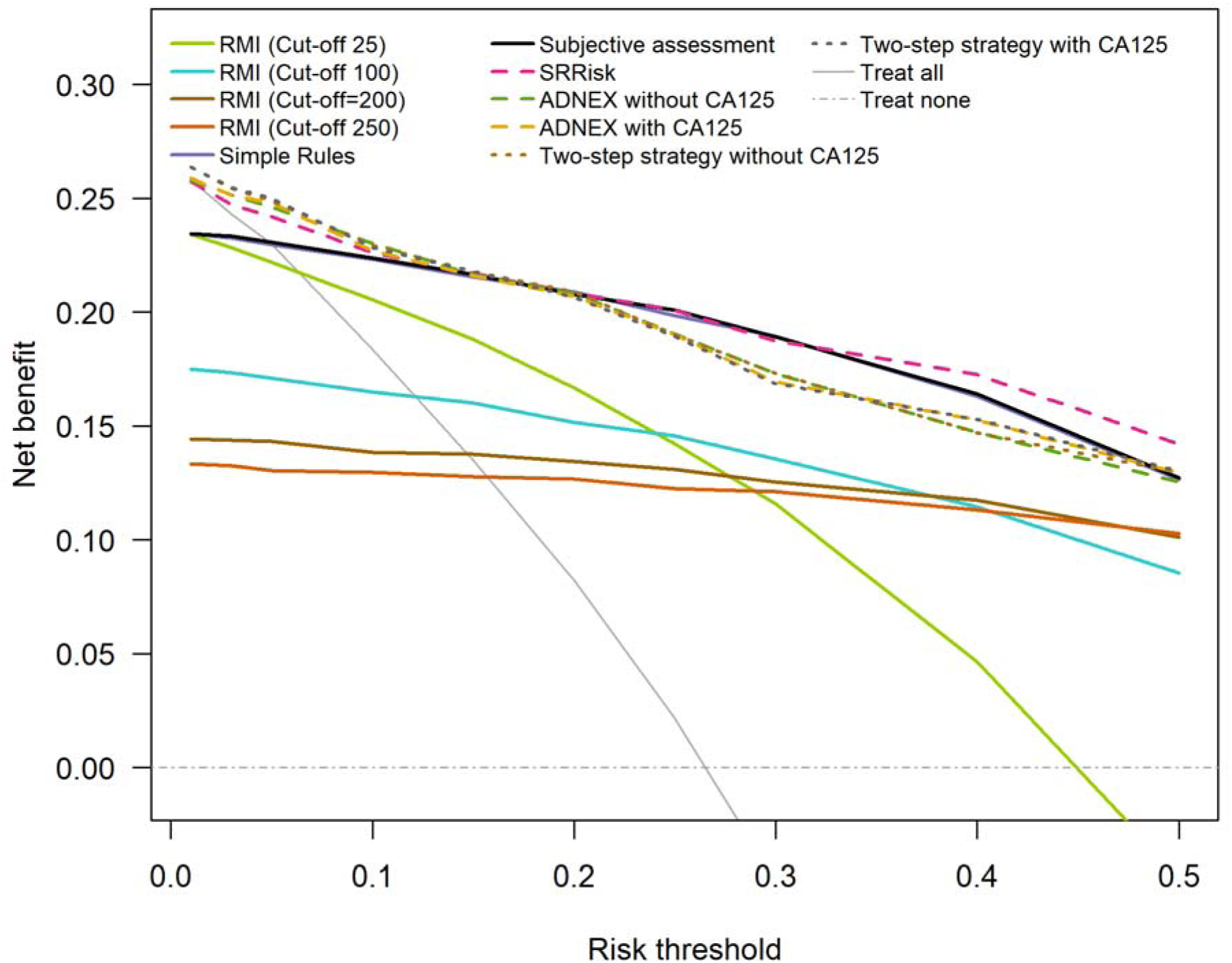
Decision curves for risk models, Risk of Malignancy Index (RMI), Simple Rules and subjective assessment based on meta-analysis of data from 21 centers. The curves show net benefit at several thresholds between 1% and 50%. A model is clinically useful if it is superior to both treat all and treat none. At risk thresholds below 15%, using RMI at cutoff 200 is worse than treating everyone (i.e.,worse than referring all women with an adnexal mass to a gynecological oncology center).

The ability of ADNEX and the two-step strategy to discriminate between different tumor types is shown in Table 3 and Supplementary Table S8. The PDI ranged from 0.44 to 0.55 for all four models. ADNEX and the two-step strategy had the same ability to discriminate between the five tumor types. Performance was poorest for discrimination between Stage I primary invasive tumors and metastases (AUROC 0.70 for all four models) and between Stage II-IV primary invasive tumors and metastases (AUROC 0.59 for models without Ca125 and 0.74 for models with CA125). AUROCs ranged from 0.93 to 0.98 for distinguishing benign tumors from invasive malignancies (primary or metastastes). Calibration of the estimated risks for the five tumor types was similar for ADNEX and the two-step strategy irrespective of whether or not CA125 was included as a predictor (Supplementary Table S9). All four models underestimated the risk of borderline, stage I primary invasive, and secondary metastatic tumors (O:E >1) but overestimated the likelihood of a stage II-IV ovarian malignancy and benign tumor (O:E<1).

**Table 3.** Polytomous discrimination index (PDI) of Assessment of Different NEoplasias in the adneXa (ADNEX), and of the two-step strategy (pooled analysis).

|  | **PDI (95% CI)** |
| --- | --- |
| ADNEX without CA125 | 0.49 (0.47; 0.53) |
| ADNEX with CA125 | 0.55 (0.51; 0.59) |
| Two-step strategy without CA125 | 0.49 (0.47; 0.52) |
| Two-step strategy with CA125 | 0.55 (0.51; 0.59) |

### Subgroup analyses

Tumor outcome and percentage of missing CA125 values in subgroups according to menopausal status, type of center and ultrasound examinerś level of experience are shown in Table 4. In all subgroups the AUROCs were higher for the IOTA models than for RMI (Figure 5). The AUROCs of the IOTA models were ≥0.90 in all subgroups (0.90 – 0.95). They were slightly higher in pre-than post-menopausal patients, in non-oncology than oncology centers, and for EFSUMB Level 3 ultrasound examiners than for EFSUMB Level 1 or 2 ultrasound examiners. The sensitivity of subjective assessment was higher for level 3 and level 2 examiners than for level 1 examiners (0.96 vs 0.92 vs 0.86), but the corresponding specificity was lower (0.84 vs 0.87 vs 0.97) (Supplementary Table S10). The sensitivity of Simple Rules (inconclusive cases classified as malignant) was higher for level 3 than level 2 and 1 examiners (0.95 vs 0.88 vs 0.82) with the corresponding specificity being lower (0.80 vs 0.85 vs 0.92).

**Figure 5.**
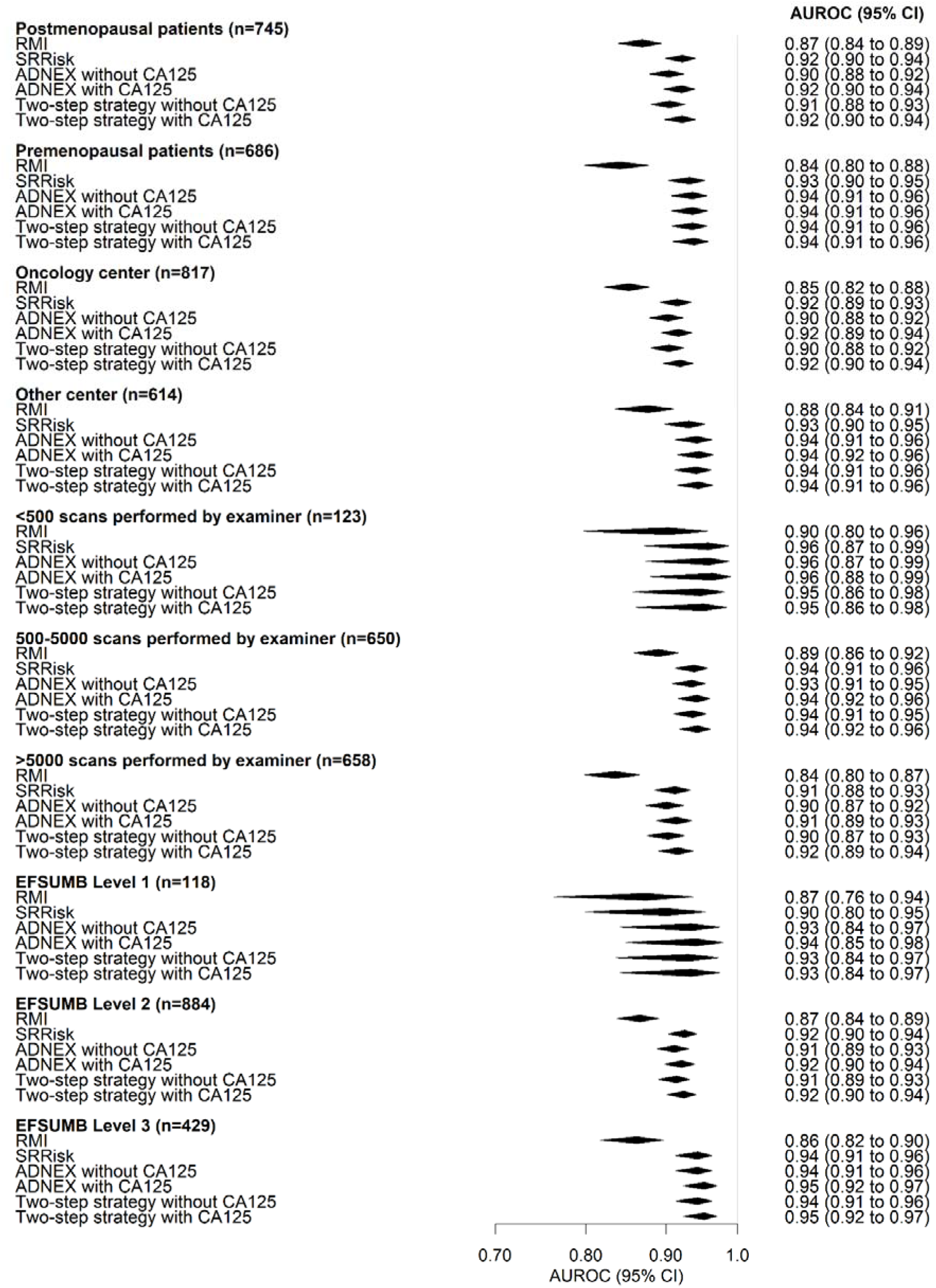
Forest plot of area under the receiver operating characteristic curve (AUROC) for prespecified subgroups (pooled data). CI; Confidence interval, PI; Predicition interval, EFSUMB; European Federation of Societies for Ultrasound in Medicine and Biology.

**Table 4.** Tumor outcome and percentage of missing CA 125 values for all pre-specified subgroups.

| Subgroup | N | Outcome |  | Missing CA125 |
| --- | --- | --- | --- | --- |
|  |  | Benign | Malignant |  |
| Postmenopausal | 745 | 444 (60) | 301 (40) | 176 (24) |
| Premenopausal | 686 | 551 (80) | 135 (20) | 218 (32) |
| Oncology center | 817 | 583 (64) | 331 (36) | 219 (27) |
| Other center | 614 | 412 (80) | 105 (20) | 175 (28) |
| Level of experience of ultrasound examiners |  |  |  |  |
| <500 scans | 123 | 102 (83) | 21 (17) | 41 (33) |
| 500-5000 scans | 650 | 476 (73) | 174 (27) | 108 (17) |
| >5000 scans | 658 | 417 (63) | 241 (37) | 245 (37) |
| EFSUMB level of ultrasound examiners |  |  |  |  |
| Level 1 | 118 | 96 (81) | 22 (19) | 38 (32) |
| Level2 | 884 | 605 (68) | 279 (32) | 248 (28) |
| Level 3 | 429 | 294 (69) | 135 (31) | 108 (25) |

Results are shown as n (%)
EFSUMB; European Federation of Societies for Ultrasound in Medicine and Biology

Calibration of the IOTA models in the subgroups is shown in Supplementary Figure S8. In all subgroups, the malignancy risk tended to be underestimated (point estimate for O:E ratio >1) by the IOTA models, with less underestimation of malignancy risk in postmenopausal than premenopausal patients and in oncology than non-oncology centers. In all subgroups, SRRisk had the least understimation. The 95% CIs for O:E ratios were very wide for EFSUMB level 1 examiners and for examiners that had performed <500 scans at study start due to the small sample size for these groups.

## Discussion

### Principal findings

SRRisk, ADNEX and the IOTA two-step strategy (with or without CA125) discriminated well between being and malignant adnexal masses and were superior to RMI when validated on a national basis in 21 Italian centers by ultrasound examiners with different levels of ultrasound experience. All IOTA methods had higher net benefit than RMI. SSRisk, ADNEX and the two step-strategy (with or without CA125) had the highest net benefit at risk thesholds below 15%.

### Strengths and limitations

Our study is the first prospective national multicenter study to validate IOTA models and to validate them in the hands of ultrasound examiners with different levels of experience. We assessed the diagnostic performance both in terms of discrimination, calibration and clinical utility. Limitations are the small number of EFSUMB level 1 examiners and examiners that had perfomed <500 scans at the start of the study, the small number of centers from the south of Italy (our aim was to include centers homogenously distributed all over Italy), and that CA125 was missing in a substantial proportion of patients (28%). We adressed the missing CA125 values using multiple imputation. Using histology as reference standard can be seen both as a strength and as a limitation. The strength is that using the same reference standard in all patients avoids differential verification bias.The limitation is that our results might not be applicable to all adnexal masses, which include also those managed with clinical and ultrasound follow-up.

### Comparison with other studies

The results of our study agree well with those in other validation studies.^9,17,23,31,48–52^ The discriminative performance of ADNEX in our study (AUROC 0.92 and 0.91 for ADNEX with and without CA125, respectively) was similar to that reported in a meta-analysis including 17007 adnexal masses examined with ultrasound in different countries and settings in 47 studies (AUROC 0.93 both for ADNEX with and without CA125). ^48^ It was only slightly poorer than that in a large international multicenter study conducted by the IOTA group (AUROC 0.94 both for ADNEX with and without CA125) and in a large single center study conducted in a private center in Barcelona, Spain (AUROC 0.95). ^22,52^ The discriminative performance of the two-step stragey was also slightly poorer than in two other large studies ^17,52^ (AUROC 0.92 vs 0.95 when ADNEX with CA125 was used as second step test; AUROC 0.91 vs 0.94 vs. 0.95 when ADNEX without CA125 was used as second step test). Whether the small differences in discriminative performance are explained by differences in tumor characteristics (the studies cited included also patients managed expectantly) or in ultrasound expertise is difficult to know. We found the discriminative ability and the clinical utility of ADNEX, the two-step strategy and SRRisk to be superior to those of RMI, which agrees with the results of other studies. ^17,22^ Both in our study and in others, the IOTA models underestimated the risk of malignancy, the best calibrated model being SRRisk, and the models being better calibrated in postmenopausal than premenopusal patients. ^17,22^ The Benign Descriptors were applicable in a lower proportion of patients in our study than in those by Landolfo et al and Pascual et al (23% vs 37% vs 77%). ^17,52^ This is explained by the the other two studies including also patients managed expectantly and by the study by Pascual et al being an extreme low-risk population. ^17,52^ The classification performance of the Simple Rules (inconclusive cases classified as malignant) in our study (sensitivity 0.90, specificity 0.85) is reasonably similar to that reported in published meta-analyses (sensitivity and specificity 0.93 and 0.81,^50^ sensitivity and specificity 0.94 and 0.76, ^51^ sensitivity and specificity 0.93 and 0.80 ^9^) and in the international multicenter study by van Calster et al (sensitivity and specificity 0.90 and 0.87 ^22^).

We found the sensitivity of subjective assessment to be 0.93 and the specificity to be 0.88, which is almost identical to the sensitivity and specificity of subjective assessment reported in a meta-analysis (sensitivity 0.93, specificity 0.89).^9^ The classification performance of subjective assessment is heavily dependent on the experience of the ultrasound examiner.^53^ The performance of risk calculation models should be less dependent on ultrasound skill as long as the ultrasound examiner is familiar with the definitions of the variables in the models. Nonetheless, we found some small differences in the discriminative and calibration performance of the IOTA models between examiners with different levels of experience, with performance being slightly better in the group of EFSUMB level 3 examiners. However, it is difficult to interpret these differences, because they may be explained by a difference in tumor types between the groups.

### Implications in clinical practice

The good performance of the IOTA models in our study, which includes also ultrasound examiners with little ultrasound experience and both local, regional and university hospitals, supports that IOTA models can be widely applied in clinical practice. Our findings also support the recommendation by Landolfo et. al ^17^ to use the two-step strategy. The two-step strategy had almost the same discrimination and calibration performance and almost the same clinical utility as ADNEX at risk thresholds up to 20% (and better clinical utility than ADNEX at the lowest risk thresholds) in our study, but the two-step strategy is easier to use than ADNEX while still offering the advantage of providing an estimate of the likelihood of four types of malignancy.

### Future perspectives

Prospective studies including a very large number of ultrasound examiners with limited experience are needed to confirm our results. It would be important to investigate the effect of using the IOTA models in impact studies. ^54^ Such studies will show whether use of IOTA models in daily practice improve decision making and, ultimately, patient outcome.

### Conclusions

SRRisk, ADNEX and the two step strategy with or without CA125 had similar and good ability to distinguish benign from malignant adnexal tumours in patients examined by either expert or non-expert ultrasound operators in Italy, and they were all superior to RMI. Our results support the recommendation by the IOTA-group to use the two-step strategy to characterize ovarian tumors.

## Data Availability

All data produced in the present study are available upon reasonable request to the authors

