## Supplementary materials for "External validation of ultrasound-based models for discrimination between benign and malignant adnexal masses in Italy: the prospective multicenter IOTA phase 6 study"

**Supplementary Table S1**. Tumor outcome according to center.

|  |  |  |  | **Tumor outcome** | | | | | |
| --- | --- | --- | --- | --- | --- | --- | --- | --- | --- |
| **Center** | **Oncological**  **center** | **Hospital** | **N (%)** | **Benign**  **n (%)** | **Borderline n (%)** | **Stage I n (%)** | **Stage II-IV**  **n (%)** | | **Metastatic n (%)** |
| Varese | No | University | 251 (18) | 199 (79) | 7 (3) | 18^a^ (7) | 20 (8) | 7 (3) | |
| Brescia | Yes | Regional | 194 (14) | 123 (64) | 22 (11) | 20 (10) | 16 (8) | 13 (7) | |
| Pisa | Yes | University | 151 (11) | 88 (58) | 19 (13) | 17 (11) | 23 (15) | 4 (3) | |
| Bari A | Yes | University | 93 (6) | 66 (71) | 2 (2) | 5^a^ (5) | 17 (18) | 3 (3) | |
| Trento | Yes | Regional | 85 (6) | 62 (73) | 8 (9) | 4 (5) | 7 (8) | 4 (5) | |
| Pavia | Yes | University | 79 (5) | 47 (59) | 5 (10) | 11 (22) | 10 (20) | 6 (12) | |
| Cuneo | Yes | Regional | 77 (5) | 29 (38) | 8 (10) | 14 (18) | 16 (21) | 10 (13) | |
| Messina | No | University | 61 (4) | 58 (95) | 3 (5) | 0 | 0 | 0 | |
| Santorso | No | Local | 56 (4) | 41 (73) | 4 (7) | 3^a^ (5) | 6 (11) | 2 (4) | |
| Torino | Yes | Regional | 52 (4) | 31 (60) | 8 (15) | 4 (8) | 6 (11) | 3 (6) | |
| Carpi | No | Local | 51 (4) | 47 (92) | 3 (6) | 1 (2) | 0 | 0 | |
| Verona | No | Regional | 46 (3) | 40 (87) | 1 (2) | 0 | 4 (9) | 1 (2) | |
| Foggia | No | University | 46(3) | 32 (70) | 8 (17) | 1 (2) | 5 (11) | 0 | |
| Bari B | Yes | Regional | 38 (3) | 23 (60) | 4 (11) | 4 (11) | 5 (13) | 2 (5) | |
| Treviso | No | Regional | 34 (2) | 23 (68) | 1 (3) | 2 (6) | 8 (23) | 0 | |
| Asti | Yes | Regional | 31 (2) | 14 (45) | 3 (10) | 4 (13) | 9 (29) | 1 (3) | |
| Firenze | No | Regional | 20 (1) | 15 (75) | 3 (15) | 0 | 1 (5) | 1 (5) | |
| Padova | No | Regional | 20 (1) | 18 (90) | 2 (10) | 0 | 0 | 0 | |
| Roma | No | Local | 18 (1) | 16 (89) | 2 (11) | 0 | 0 | 0 | |
| Bolzano | Yes | Regional | 17 (1) | 13 (76) | 2 (12) | 1 (6) | 1 (6) | 0 | |
| Montebelluna | No | Local | 11 (1) | 10 (91) | 0 | 0 | 0 | 1 (9) | |
| Oncological center |  |  | 817 (57) | 583 (64) | 85 (9) | 85 (9) | 114 (12) | 47 (5) | |
| Non oncological center |  |  | 614 (43) | 412 (80) | 30 (6) | 24 (5) | 40 (8) | 11 (2) | |

^a^ Including 1 with unknown FIGO stage

For tumor outcome, percentages are calculated per row

**Supplementary table S2.** Specific histological diagnosis.

|  | *n (%)* |
| --- | --- |
| Histological diagnosis: Benign | 930 (65) |
| Serous cystadenofibroma/ serous cystadenoma | 255 (18) |
| Teratoma | 163 (11) |
| Endometrioma | 140 (10) |
| Mucinous cystadenofibroma/ Mucinous cystadenoma | 133 (9) |
| Simple cyst/para-ovarian or salpingeal cyst | 88 (6) |
| Fibroma | 84 (6) |
| Other | 21 (1) |
| Thecoma | 19 (1) |
| Haemorragic corpus luteum cyst | 10 (1) |
| Normal | 6 (<1) |
| Functional cyst | 4 (<1) |
| Peritoneal pseudocyst | 4 (<1) |
| Inclusion cyst | 3 (<1) |
| Histological diagnosis: Rare benign | 26 (2) |
| Brenner | 12 (1) |
| Struma ovarii | 9 (1) |
| Other | 3 (<1) |
| Leydig | 2 (<1) |
| Histological diagnosis: Infectious acute chronic | 34 (2) |
| Hydrosalpinx | 12 (1) |
| Abscess | 11 (1) |
| Salpingitis | 8 (1) |
| Other | 3 (<1) |
| Histological diagnosis: Uterine | 5 (<1) |
| Fibroid | 3 (<1) |
| Subserous adenomyoma | 1 (<1) |
| Other | 1 (<1) |
| Histological diagnosis: Borderline | 115 (8) |
| Serous | 62 (4) |
| Mucinous gastrointestinal | 36 (2) |
| Mucinous endocervical | 10 (1) |
| Other | 7 (<1) |
| Histological diagnosis: Malignant | 260 (18) |
| Epithelial | 230 (16) |
| Stromal | 15 (1) |
| Tubal | 11 (1) |
| Germ cells | 4 (<1) |
| Histological diagnosis: Metastatic | 50 (3) |
| Gastrointestinal | 22 (1) |
| Other | 17 (1) |
| Breast | 5 (<1) |
| Krukenberg | 4 (<1) |
| Lymphoma | 2 (<1) |
| Histological diagnosis: Exceptional | 11 (1) |

**Supplementary Table S3.** Comparison of the current dataset with development datasets of the models assessed.

| **Variable** | **Model** | **RMI**  **development (n=143, 1**  **center)** ^1^ | **SR**  **development (n=1066,**  **9 centers)** ^2^ | **SRRisk development (n=4848,**  **22 centers)** ^3^ | **ADNEX**  **development (n=5914,**  **24 centers)** ^4^ | **Two-step strategy**  **(n=4905,**  **17 centers)**^5^ | **Current data**  **(n=1431,**  **21 centers)** |
| --- | --- | --- | --- | --- | --- | --- | --- |
| Benign outcome, % |  | 71 | 75 | 66 | 67 | 70 | 70 |
| Malignant outcome, % |  | 29 | 25 | 34 | 33 | 20 | 30 |
| Borderline outcome, % |  | - | 5 | 6 | 6 | 4 | 8 |
| Stage I outcome, % |  | - | 5 | 6 | 6 | 4 | 8 |
| Stage II-IV outcome, % |  | - | 11 | 18 | 17 | 9 | 11 |
| Metastatic outcome, % |  | - | 4 | 4 | 4 | 3 | 4 |
| Uncertain outcome, % |  | 0 | 0 | 0 | 0 | 10 | 0 |
| Age (years), mean | ADNEX | 52 | 47 | 48 | 48 | 49 | 50 |
| Postmenopausal, % | RMI | 58 | 41 | 41 | 41 | 44 | 52 |
| CA125 (U/ml), mean, when available | RMI, ADNEX | 48 | 305 | 363 | 352 | 318 | 166 |
| CA125 (U/ml), median, when available | RMI, ADNEX | - | 23 | 33 | 30 | 25 | 19 |
| CA125, % missing | RMI, ADNEX | 0 | 24 | 32 | 31 | 53 | 27 |
| Max. diameter of lesion (mm), median | ADNEX, 2-step | - | 68 | 69 | 69 | 55 | 69 |
| Proportion solid tissue, median | ADNEX | - | 0.06 | 0.13 | 0.11 | 0 | 0.05 |
| Presence of solid areas, % | RMI | - | 52 | 53 | 53 | 35 | 51 |
| Irregular internal cyst walls, % | SR, SRRisk | - | 45 | 39 | 40 | 31 | 39 |
| Acoustic shadows, % | SR, SRRisk, ADNEX, 2-step | - | 10 | 13 | 13 | 15 | 23 |
| Ascites, % | RMI, SR, SRRisk, ADNEX | - | 13 | 12 | 12 | 6 | 8 |
| Number of papillations, mean | ADNEX | - | 0.60 | 0.41 | 0.45 | 0.25 | 0.56 |
| Bilateral, % | RMI | - | 20 | 19 | 19 | 17 | 20 |
| Multilocular cyst, % | RMI | - | 45 | 37 | 39 | 34 | 34 |
| >10 locules, % | ADNEX | - | 9 | 8 | 8 | 8 | 11 |
| Abdominal metastases, % | RMI | - | - | 14 | 14 | 7 | 7 |
| Unilocular cyst, % | SR, SRRisk, 2-step | - | 29 | 30 | 30 | 44 | 28 |
| Solid areas, but smaller than 7mm, % | SR, SRRisk | - | 4 | 2 | 2 | 2 | 4 |
| Smooth multilocular cyst  <100mm, % | SR, SRRisk | - | 11 | 10 | 10 | 13 | 13 |
| No blood flow (color score 1), % | SR, SRRisk | - | 22 | 29 | 28 | 41 | 51 |
| Irregular solid tumor, % | SR, SRRisk | - | 6 | 6 | 6 | 4 | 7 |
| At least 4 papillations, % | SR, SRRisk | - | 9 | 5 | 6 | 3 | 8 |
| Irregular multilocular-solid cyst  ≥100mm, % | SR, SRRisk | - | 9 | 8 | 8 | 4 | 9 |
| Very strong blood flow (color score 4), % | SR, SRRisk | - | 14 | 12 | 12 | 9 | 13 |
| Unilocular, ground glass echogenicity, premenopausal, <100mm, % | 2-step | - | 9 | 10 | 9 | 10 | 6 |
| Unilocular, mixed echogenicity, acoustic shadows, premenopausal, <100mm, % | 2-step | - | 2 | 3 | 3 | 4 | 3 |
| Unilocular, anechoic cyst fluid, smooth, <100mm, % | 2-step | - | 7 | 6 | 6 | 14 | 6 |
| All other unilocular cyst, smooth, <100mm, % | 2-step | - | 5 | 5 | 5 | 8 | 7 |

Max, maximum; RMI, risk of malignancy index; SR, Simple Rules; SRRisk, Simple Rules risk model; ADNEX, Assessment of Different NEoplasias in the adneXa.; 2-step, two-step strategy. RMI development.

**Supplementary Table S4**. Outcome for tumors to which a benign descriptor (BD) applied (pooled data).

|  | | **Tumor subtype** | | | | |
| --- | --- | --- | --- | --- | --- | --- |
| **Benign descriptor** | **N** | **Benign**  **n (%)** | **Borderline**  **n (%)** | **Stage I invasive**  **n (%)** | **Stage II-IV invasive**  **n (%)** | **Secondary metastatic**  **n (%)** |
| Any descriptor | 328 | 325  (99) | 3  (1) | 0 | 0 | 0 |
| BD 1 | 84 | 83  (99) | 1  (1) | 0 | 0 | 0 |
| BD 2 | 50 | 50  (100) | 0 | 0 | 0 | 0 |
| BD 3 | 89 | 89  (100) | 0 | 0 | 0 | 0 |
| BD 4 | 105 | 103  (98) | 2  (2) | 0 | 0 | 0 |

The benign simple descriptors applied to 328/1431 (23%) patients. Percentages are calculated per row.

**Supplementary Table S5**. Sensitivity, specificity, negative predictive value (NPV), and positive predictive value (PPV) of risk models at pre-specified risk of malignancy thresholds based on meta-analysis of results from 21 centers.

| **Risk threshold** | **Model** | **Sensitivity (95% CI)** | **Specificity (95% CI)** | **NPV (95% CI)** | **PPV (95% CI)** |
| --- | --- | --- | --- | --- | --- |
| 1% | SRRisk | 0.98 (0.95; 0.99) | 0.53 (0.44; 0.61) | 0.98 (0.96; 0.99) | 0.45 (0.38; 0.52) |
|  | ADNEX without CA125 | >0.99 (0.98; >0.99) | 0.16 (0.12; 0.20) | 0.99 (0.96; >0.99)^C,A^ | 0.31 (0.24; 0.38) |
|  | ADNEX with CA125 | >0.99 (0.98; >0.99) | 0.16 (0.12; 0.21) | 0.99 (0.96; >0.99)^C,A^ | 0.31 (0.25; 0.38) |
|  | Two-step without CA125 | 0.99 (0.98; >0.99) | 0.37 (0.29; 0.45) | 0.99 (0.97; >0.99)^A^ | 0.39 (0.33; 0.45) |
|  | Two-step with CA125 | 0.99 (0.98; >0.99) | 0.37 (0.29; 0.46) | 0.99 (0.97; >0.99)^A^ | 0.39 (0.33; 0.46) |
| 3% | SRRisk | 0.96 (0.92; 0.98) | 0.67 (0.59; 0.74) | 0.98 (0.95; 0.99) | 0.54 (0.49; 0.59) |
|  | Adnex without CA125 | >0.99 (0.94; >0.99) | 0.46 (0.32; 0.60) | 0.99 (0.94; >0.99)^C^ | 0.44 (0.39; 0.49) |
|  | ADNEX with CA125 | 0.99 (0.94; >0.99) | 0.51 (0.39; 0.64) | 0.99 (0.95; >0.99) | 0.47 (0.42; 0.51) |
|  | Two-step without CA125 | 0.99 (0.99; 0.99) | 0.54 (0.42; 0.64) | 0.99 (0.95; >0.99) | 0.48 (0.43; 0.53) |
|  | Two-step with CA125 | 0.99 (0.99; >0.99) | 0.57 (0.46; 0.68) | 0.99 (0.96;>0.99) | 0.50 (0.45; 0.54) |
| 5% | SRRisk | 0.96 (0.91; 0.98) | 0.69 (0.61; 0.76) | 0.97 (0.95; 0.99) | 0.55 (0.50; 0.60) |
|  | ADNEX without CA125 | 0.98 (0.92; 0.99) | 0.64 (0.53; 0.75) | 0.98 (0.95; 0.99) | 0.53 (0.50; 0.57) |
|  | ADNEX with CA125 | 0.97 (0.93; 0.99) | 0.68 (0.57; 0.77) | 0.98 (0.96; 0.99) | 0.56 (0.52; 0.59) |
|  | Two-step witout CA125 | 0.98 (0.92; 0.99) | 0.66 (0.55; 0.75) | 0.98 (0.96; 0.999) | 0.54 (0.50; 0.58) |
|  | Two-step with CA125 | 0.97 (0.93; 0.99) | 0.69 (0.58; 0.78) | 0.98 (0.96; 0.99) | 0.56 (0.53; 0.60) |
| 10% | SRRisk | 0.93 (0.88; 0.96) | 0.78 (0.69; 0.85) | 0.96 (0.94; 0.98) | 0.61 (0.58; 0.65) |
|  | ADNEX without CA125 | 0.94 (0.88; 0.97) | 0.77 (0.66; 0.85) | 0.96 (0.94; 0.98) | 0.61 (0.57; 0.65) |
|  | ADNEX with CA125 | 0.92 (0.87; 0.95) | 0.80 (0.71; 0.86) | 0.96 (0.93; 0.97) | 0.64 (0.60; 0.68) |
|  | Two-step without CA125 | 0.94 (0.88; 0.97) | 0.77 (0.66; 0.85) | 0.96 (0.94; 0.98) | 0.61 (0.57; 0.65) |
|  | Two-step with CA125 | 0.92 (0.87; 0.95) | 0.80 (0.71; 0.86) | 0.96 (0.93; 0.97) | 0.64 (0.60; 0.68) |
| 15% | SRRisk | 0.91 (0.85; 0.94) | 0.83 (0.76; 0.89) | 0.95 (0.93; 0.97) | 0.67 (0.63; 0.71) |
|  | ADNEX without CA125 | 0.90 (0.84; 0.94) | 0.83 (0.76; 0.89) | 0.95 (0.93; 0.97) | 0.68 (0.64; 0.71) |
|  | ADNEX with CA125 | 0.89 (0.84; 0.93) | 0.85 (0.78; 0.89) | 0.95 (0.92; 0.96) | 0.70 (0.66; 0.74) |
|  | Two-step without CA125 | 0.90 (0.84; 0.94) | 0.83 (0.76; 0.89) | 0.95 (0.93; 0.97) | 0.68 (0.64; 0.71) |
|  | Two-step with CA125 | 0.89 (0.84; 0.93) | 0.85 (0.78; 0.89) | 0.95 (0.92; 0.96) | 0.70 (0.66; 0.74) |
| 20% | SRRisk | 0.89 (0.84; 0.92) | 0.86 (0.81; 0.90) | 0.95 (0.92; 0.96) | 0.72 (0.67; 0.77) |
|  | ADNEX without CA125 | 0.88 (0.82; 0.92) | 0.87 (0.80; 0.92) | 0.94 (0.92; 0.96) | 0.71 (0.67; 0.75) |
|  | ADNEX with CA125 | 0.87 (0.82; 0.91) | 0.90 (0.84; 0.93) | 0.94 (0.92; 0.96) | 0.75 (0.71; 0.79) |
|  | Two-step without CA125 | 0.88 (0.82; 0.92) | 0.87 (0.80; 0.92) | 0.94 (0.92; 0.96) | 0.71 (0.67; 0.75) |
|  | Two-step with CA125 | 0.87 (0.82; 0.91) | 0.90 (0.84; 0.93) | 0.94 (0.92; 0.96) | 0.75 (0.71; 0.79) |
| 25% | SRRisk | 0.89 (0.84; 0;92) | 0.86 (0.81; 0.90) | 0.95 (0.92; 0.96) | 0.72 (0.67; 0.77) |
|  | ADNEX without CA125 | 0.84 (0.78; 0.89) | 0.89 (0.84; 0.93) | 0.93 (0.91; 0.95) | 0.74 (0.70; 0.78) |
|  | ADNEX with CA125 | 0.82 (0.76; 0.87) | 0.91 (0.86; 0.94) | 0.93 (0.90; 0.94) | 0.77 (0.73; 0.81) |
|  | Two-step without CA125 | 0.84 (0.78; 0.89) | 0.89 (0.84; 0.93) | 0.93 (0.91; 0.95) | 0.74 (0.70; 0.78) |
|  | Two-step with CA125 | 0.82 (0.76; 0.87) | 0.91 (0.86; 0.94) | 0.93 (0.90; 0.94) | 0.77 (0.73; 0.81) |
| 30% | SRRisk | 0.84 (0.77; 0.89) | 0.91 (0.86; 0.94) | 0.93 (0.91; 0.95) | 0.77 (0.73; 0.81) |
|  | ADNEX without CA125 | 0.80 (0.72; 0.86) | 0.91 (0.86; 0.94) | 0.92 (0.89; 0.94) | 0.76 (0.72; 0.80) |
|  | ADNEX with CA125 | 0.77 (0.68; 0.83) | 0.93 (0.88; 0.96) | 0.91 (0.88; 0.93) | 0.80 (0.76; 0.84)^R^ |
|  | Two-step without CA125 | 0.80 (0.72; 0.86) | 0.91 (0.86; 0.94) | 0.92 (0.89; 0.94) | 0.76 (0.72; 0.80) |
|  | Two-step with CA125 | 0.77 (0.68; 0.83) | 0.93 (0.88; 0.96) | 0.91 (0.88; 0.93) | 0.80 (0.76; 0.84)^R^ |
| 40% | SRRisk | 0.83 (0.75; 0.88) | 0.91 (0.86; 0.95) | 0.93 (0.90; 0.95) | 0.78 (0.74; 0.82)^R^ |
|  | ADNEX without CA125 | 0.73 (0.64; 0.80) | 0.93 (0.89; 0.95) | 0.89 (0.87; 0.92) | 0.79 (0.75; 0.83) |
|  | ADNEX with CA125 | 0.73 (0.66; 0.79) | 0.94 (0.90; 0.96) | 0.89 (0.86; 0.91) | 0.82 (0.78; 0.86)^R^ |
|  | Two-step without CA125 | 0.73 (0.64; 0.80) | 0.93 (0.89; 0.95) | 0.89 (0.87; 0.92) | 0.79 (0.75; 0.83) |
|  | Two-step with CA125 | 0.73 (0.66; 0.79) | 0.94 (0.90; 0.96) | 0.89 (0.86; 0.91) | 0.82 (0.78; 0.86)^R^ |
| 50% | SRRisk | 0.73 (0.65; 0.79) | 0.93 (0.91; 0.95) | 0.90 (0.86; 0.92) | 0.82 (0.78; 0.86)^R^ |
|  | ADNEX without CA125 | 0.67 (0.59; 0.74) | 0.94 (0.91; 0.97) | 0.88 (0.85; 0.90) | 0.82 (0.77: 0.85)^R,M^ |
|  | ADNEX with CA125 | 0.67 (0.60; 0.73) | 0.95 (0.92; 0.97) | 0.87 (0.84; 0.90) | 0.84 (0.80; 0.88)^R,M^ |
|  | Two-step without CA125 | 0.67 (0.59; 0.74) | 0.94 (0.91; 0.97) | 0.88 (0.88; 0.90) | 0.82 (0.77; 0.85)^R,M^ |
|  | Two-step with CA125 | 0.67 (0.60; 0.73) | 0.95 (0.92; 0.97) | 0.87 (0.84; 0.90) | 0.84 (0.80; 0.88)^R,M^ |

R: Rome excluded from meta-analysis; M: Carpi excluded from meta-analysis; C: Cuneo excluded from meta-analysis; A: Asti excluded from meta-analysis. When a center had zero true positive (TP) and false positive (FP) at a specific threshold, it was excluded from the meta-analysis of PPV at that specific threshold. When a center had zero true negative (TN) and false negative (FN) at a specific threshold, it was excluded from the meta-analysis of NPV at that specific threshold.

**Supplementary Table S6**. Sensitivity, specificity, negative predictive value (NPV), and positive predictive value (PPV) of Risk of Malignancy Index (RMI) at pre-specified thresholds based on meta-analysis of results from 21 centers.

| **RMI threshold** | **Sensitivity (95% CI)** | **Specificity (95% CI)** | **NPV (95% CI)** | **PPV (95% CI)** |
| --- | --- | --- | --- | --- |
| 25 | 0.88 (0.84; 0.91) | 0.61 (0.54; 0.67) | 0.93 (0.89; 0.95) | 0.46 (0.38; 0.54) |
| 100 | 0.68 (0.63; 0.73) | 0.87 (0.85; 0.90) | 0.87 (0.83; 0.91) | 0.70 (0.64; 0.75) |
| 200 | 0.58 (0.52; 0.63) | 0.94 (0.92; 0.96) | 0.85 (0.80; 0.89) | 0.80 (0.76; 0.84)^P,E^ |
| 250 | 0.55 (0.48; 0.61) | 0.96 (0.94; 0.97) | 0.84 (0.79; 0.88) | 0.84 (0.79; 0.88)^P,E,T^ |

A risk threshold of 200 is often used in European guidelines ^6–9^

CI, confidence interval

P: Padova excluded from meta-analysis; E: Messina excluded from meta-analysis; T: Montebelluna excluded from meta-analysis. When a center had zero true positive (TP) and false positive (FP) at a specific threshold, it was excluded from the meta-analysis of PPV at that specific threshold. When a center had zero true negative (TN) and false negative (FN) at a specific threshold, it was excluded from the meta-analysis of NPV at that specific threshold.

**Supplementary Table S7**. Sensitivity, specificity, negative predictive value (NPV), positive predictive value (PPV) for Simple Rules (inconclusive cases are classified as malignant) and subjective assessment based on meta-analysis of results from 21 centers.

|  | **Sensitivity (95% CI)** | **Specificity (95% CI)** | **NPV (95% CI)** | **PPV (95% CI)** |
| --- | --- | --- | --- | --- |
| Simple Rules | 0.90 (0.85; 0.93) | 0.85 (0.80; 0.88) | 0.95 (0.93; 0.97) | 0.71 (0.65; 0.76) |
| Subjective assessment | 0.93 (0.90; 0.95) | 0.88 (0.84; 0.91) | 0.97 (0.95; 0.98) | 0.76 (0.70; 0.81) |

1244/1431 (87%) tumors could be classified as benign or malignant according to the Simple Rules

CI, confidence interval

**Supplementary Table S8.** Area under the receiver operating characteristics curves (AUROC) of Assessment of Different NEoplasias in the adnexa (ADNEX) and of the two-step strategy for each pair of outcome categories (pooled data).

|  | **AUROC (95% CI)** | | | |
| --- | --- | --- | --- | --- |
| **Pair of outcome categories** | **ADNEX without CA125** | **ADNEX with CA125** | **Two-step strategy without CA125** | **Two-step strategy with CA125** |
| Benign vs Borderline | 0.87  (0.84; 0.90) | 0.88  (0.84; 0.91) | 0.88  (0.84; 0.91) | 0.88  (0.84; 0.91) |
| Benign vs Stage I invasive | 0.93  (0.90; 0.95) | 0.94  (0.91; 0.96) | 0.93  (0.90; 0.96) | 0.94  (0.91; 0.96) |
| Benign vs Stage II-IV invasive | 0.98  (0.97; 0.99) | 0.98  (0.97; 0.99) | 0.97  (0.95; 0.98) | 0.98  (0.97; 0.99) |
| Benign vs Secondary metastasis | 0.96  (0.92; 0.98) | 0.96  (0.92; 0.98) | 0.96  (0.92; 0.98) | 0.96  (0.92; 0.98) |
| Borderline vs Stage I invasive | 0.82  (0.76; 0.87) | 0.81  (0.75; 0.86) | 0.82  (0.76; 0.87) | 0.81  (0.75; 0.86) |
| Borderline vs Stage II-IV invasive | 0.90  (0.86; 0.93) | 0.93  (0.89; 0.96) | 0.90  (0.86; 0.93) | 0.93  (0.89; 0.96) |
| Borderline vs Secondary metastasis | 0.87  (0.80; 0.92) | 0.87  (0.81; 0.92) | 0.87  (0.80; 0.91) | 0.87  (0.80; 0.92) |
| Stage I invasive vs Stage II-IV invasive | 0.76  (0.69; 0.81) | 0.83  (0.77; 0.88) | 0.76  (0.69; 0.81) | 0.83  (0.77; 0.88) |
| Stage I invasive vs Secondary metastasis | 0.70  (0.62; 0.78) | 0.70  (0.61; 0.78) | 0.70  (0.62; 0.78) | 0.70  (0.61; 0.78) |
| Stage II-IV invasive vs secondary metastasis | 0.59  (0.51; 0.68) | 0.74  (0.65; 0.81) | 0.59  (0.51; 0.68) | 0.74  (0.65; 0.81) |

CI; Confidence interval.

**Supplementary Table S9**. Calibration in terms of O:E ratios for Assessment of Different Neoplasias in the adnexa (ADNEX) and for the two-step strategy for each of the five tumor types (pooled data).

|  | **O:E (95% CI)** | | | |
| --- | --- | --- | --- | --- |
| **Category** | **ADNEX without CA125** | **ADNEX with CA125** | **Two-step strategy without CA125** | **Two-step strategy with CA125** |
| Benign | 0.96  (0.92; 0.99) | 0.95  (0.90; 0.99) | 0.95  (0.92; 0.99) | 0.94  (0.90; 0.99) |
| Borderline | 1.33  (1.11; 1.58) | 1.30  (1.19; 1.43) | 1.37  (1.15; 1.64) | 1.34  (1.23; 1.47) |
| Stage I invasive | 1.41  (1.17; 1.68) | 1.42  (1.30; 1.56) | 1.42  (1.19; 1.70) | 1.44  (1.31; 1.57) |
| Stage II-IV invasive | 0.81  (0.70; 0.94) | 0.86  (0.79; 0.94) | 0.81  (0.70; 0.94) | 0.86  (0.79; 0.95) |
| Secondary metastasis | 1.54  (1.20; 1.98) | 1.49  (1.35; 1.63) | 1.54  (1.20; 1.99) | 1.49  (1.36; 1.64) |

CI, confidence interval

**Supplementary Table S10**. Classification performance of subjective assessment and Simple Rules depending on the level of experience of the ultrasound examiner (pooled data).

|  | **Sensitivity (95% CI)** | **Specificity (95% CI)** | **NPV (95% CI)** | **PPV (95% CI)** |
| --- | --- | --- | --- | --- |
| *Subjective assessment* | | | | |
| <500 scans | 0.86 (0.64; 0.97) | 0.97 (0.92; 0.99) | 0.97 (0.92; 0.99) | 0.86 (0.64; 0.97) |
| 500-5000 scans | 0.94 (0.89; 0.97) | 0.87 (0.84; 0.90) | 0.97 (0.95; 0.99) | 0.73 (0.66; 0.78) |
| >5000 scans | 0.93 (0.89; 0.96) | 0.84 (0.81; 0.88) | 0.95 (0.93; 0.97) | 0.78 (0.72; 0.82) |
| EFSUMB level of the ultrasound examiner |  |  |  |  |
| Level 1 | 0.86 (0.65; 0.97) | 0.97 (0.91; 0.99) | 0.97 (0.91; 0.99) | 0.86 (0.65; 0.97) |
| Level 2 | 0.92 (0.88; 0.95) | 0.87 (0.84; 0.90) | 0.96 (0.94; 0.97) | 0.76 (0.72; 0.81) |
| Level 3 | 0.96 (0.91; 0.98) | 0.84 (0.79; 0.88) | 0.98 (0.95; 0.99) | 0.73 (0.66; 0.80) |
| *Simple rules* | | | | |
| <500 scans | 0.90 (0.70;0.99) | 0.93 (0.86;0.97) | 0.98 (0.93;>0.99) | 0.73 (0.52;0.88) |
| 500-5000 scans | 0.90 (0.85;0.94) | 0.83 (0.80;0.87) | 0.96 (0.94;0.98) | 0.67 (0.60;0.73) |
| >5000 scans | 0.89 (0.85;0.93) | 0.82 (0.78;0.86) | 0.93 (0.90;0.95) | 0.75 (0.69;0.80) |
| EFSUMB level of the ultrasound examiner |  |  |  |  |
| Level 1 | 0.82 (0.60;0.95) | 0.92 (0.84;0.96) | 0.96 (0.89;0.99) | 0.69 (0.48;0.86) |
| Level 2 | 0.88 (0.83;0.91) | 0.85 (0.82;0.87) | 0.94 (0.91;0.96) | 0.72 (0.67;0.77) |
| Level 3 | 0.95 (0.90;0.98) | 0.80 (0.75;0.85) | 0.97 (0.94;0.99) | 0.69 (0.62;0.75) |

CI; Confidence Interval, EFSUMB; European Federation of Societies for Ultrasound in Medicine and Biology.

For Simple rules, the inconclusive cases were classified as malignant.

**Supplementary Figure S1**. Forest plot with center-specific area under the receiver operating characteristic curve (AUROC) of the Risk of Malignancy Index (RMI). CI; Confidence Interval

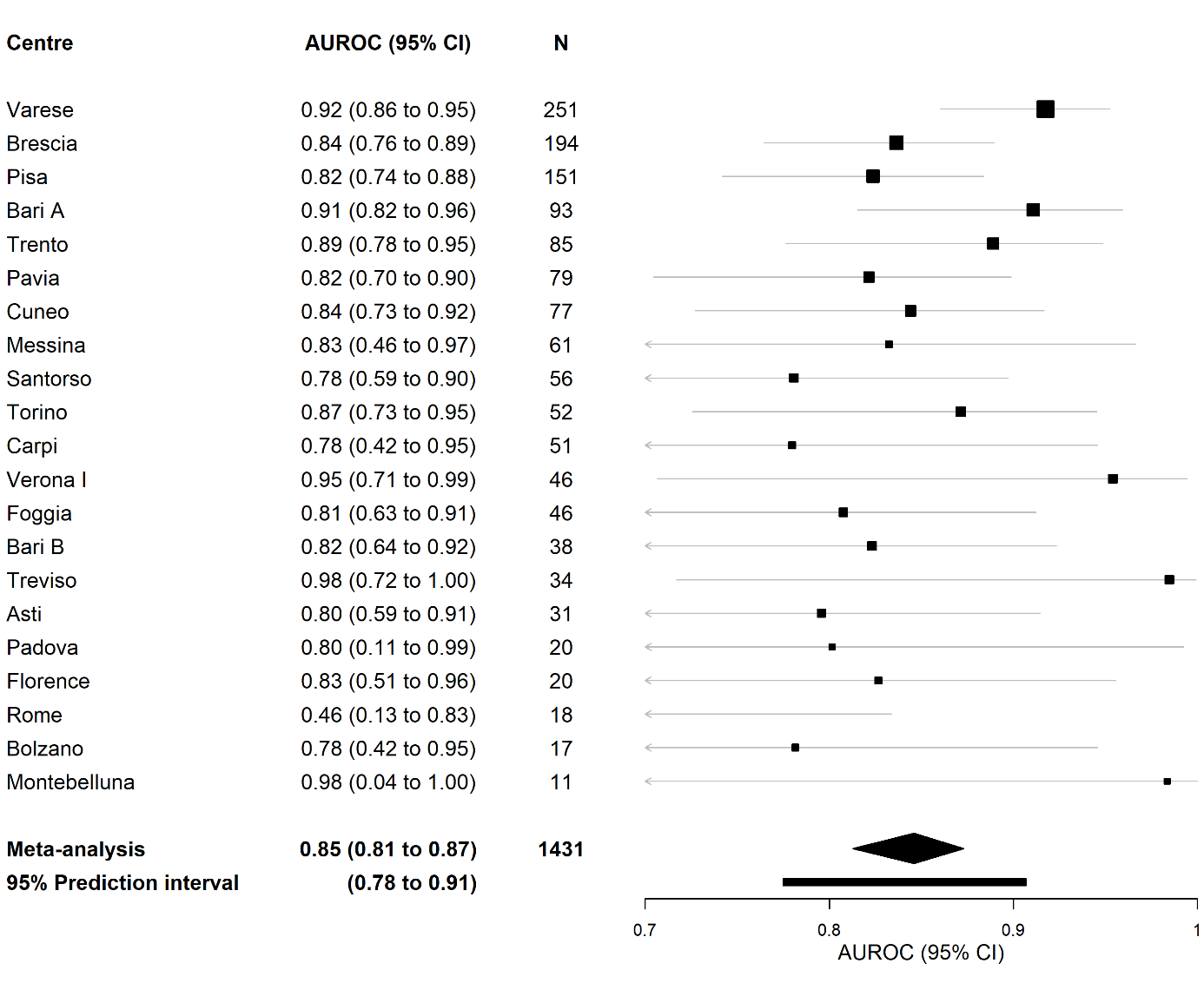

**Supplementary Figure S2**. Forest plot with center-specific area under the receiver operating characteristic curve (AUROC) of Simple Rules risk model (SRRisk). CI; Confidence Interval.

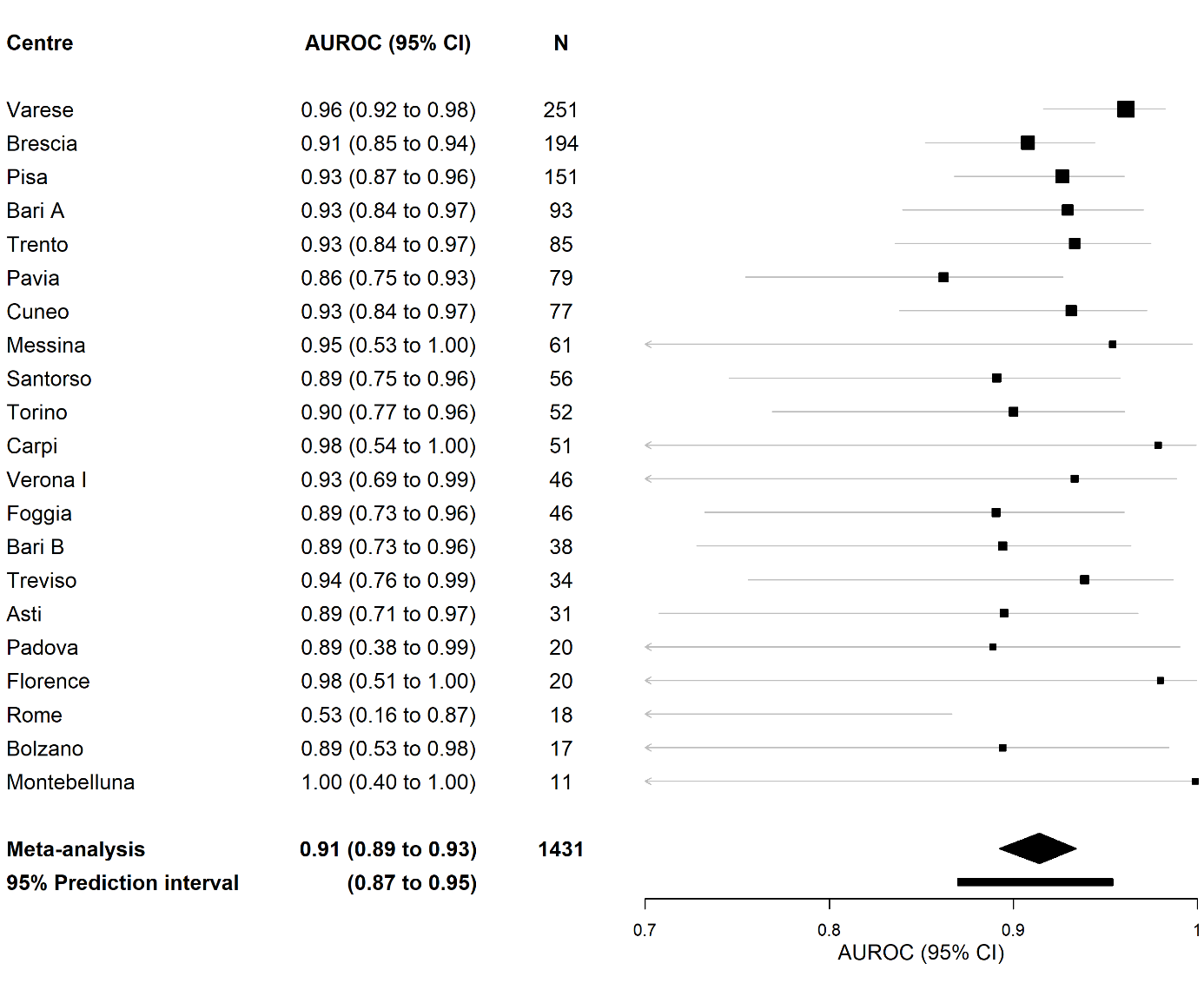

**Supplementary Figure S3**. Forest plot with center-specific area under the receiver operating characteristic curve (AUROC) of Assessment of Different Neoplasias in the adnexa (ADNEX) without CA125. For Rome, 2 tumors were borderline tumors and the rest were benign. The risk of malignancy estimated by ADNEX without CA125 was 1.95% and 1.90% for the borderline tumors. CI; Confidence Interval.

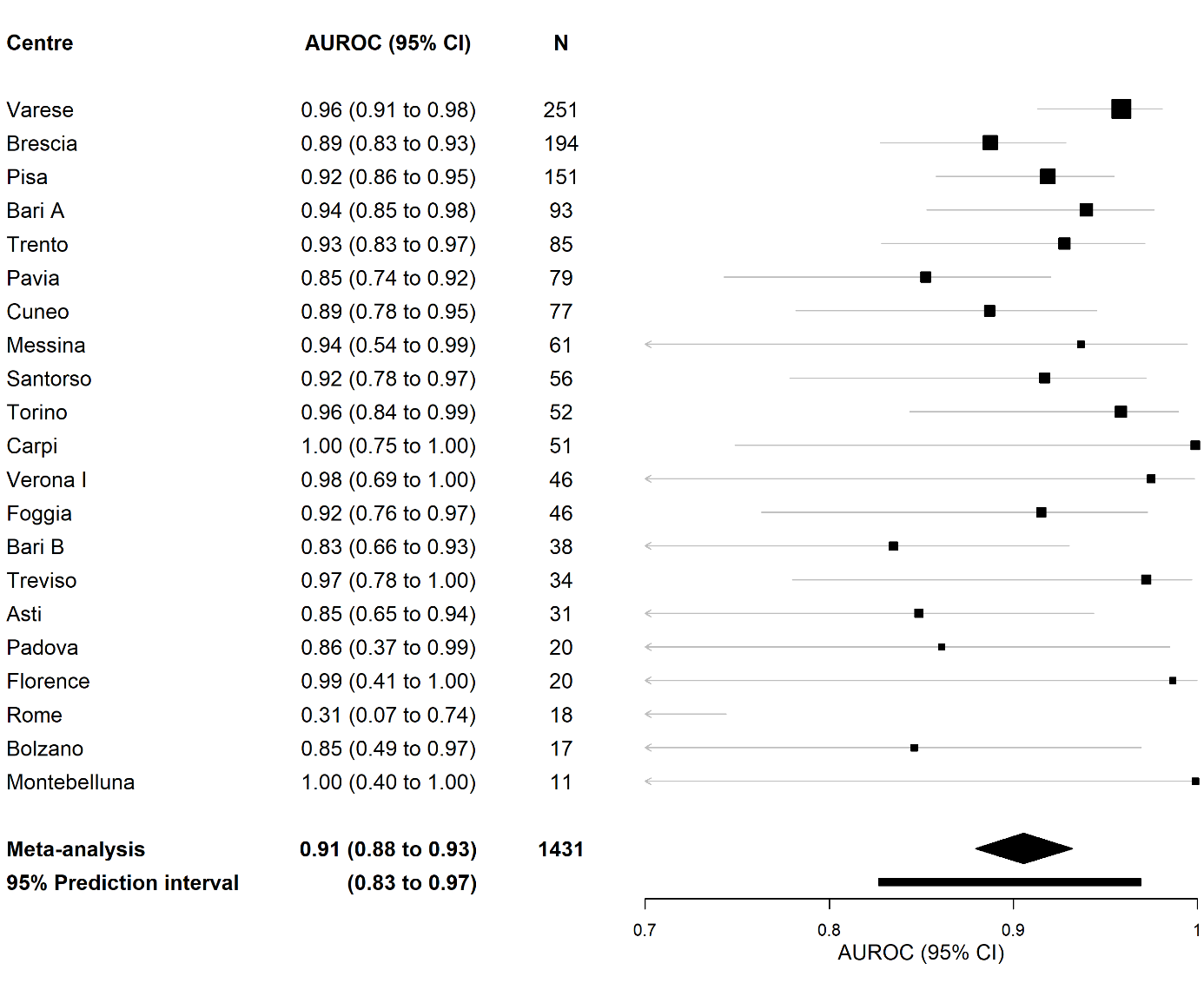

**Supplementary Figure S4**. Forest plot with center-specific area under the receiver operating characteristic curve (AUROC) of Assessment of Different Neoplasias in the adnexa (ADNEX) with CA125. For Rome, 2 tumors were borderline tumors and the rest were benign. The risk of malignancy estimated by ADNEX with CA125 was 1.83% and 1.65% for the borderline tumors. CI; Confidence Interval.

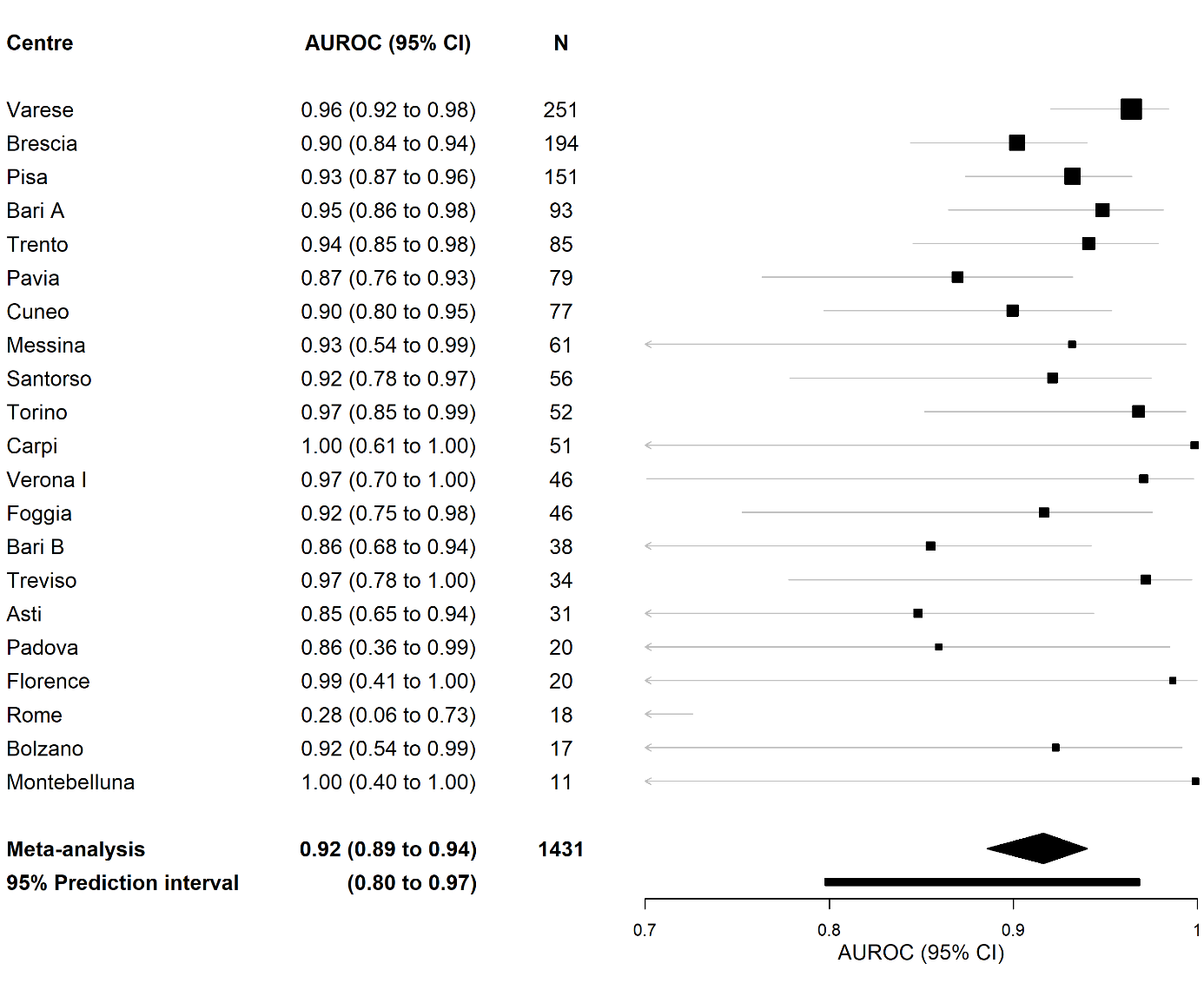

**Supplementary Figure S5**. Forest plot with center-specific area under the receiver operating characteristic curve (AUROC) of two-step strategy without CA125. For Rome, 2 tumors were borderline tumors and the rest were benign. The risk of malignancy estimated by the two-step strategy without CA125 was 1.95% and 1.90% for the borderline tumors. CI; Confidence Interval.

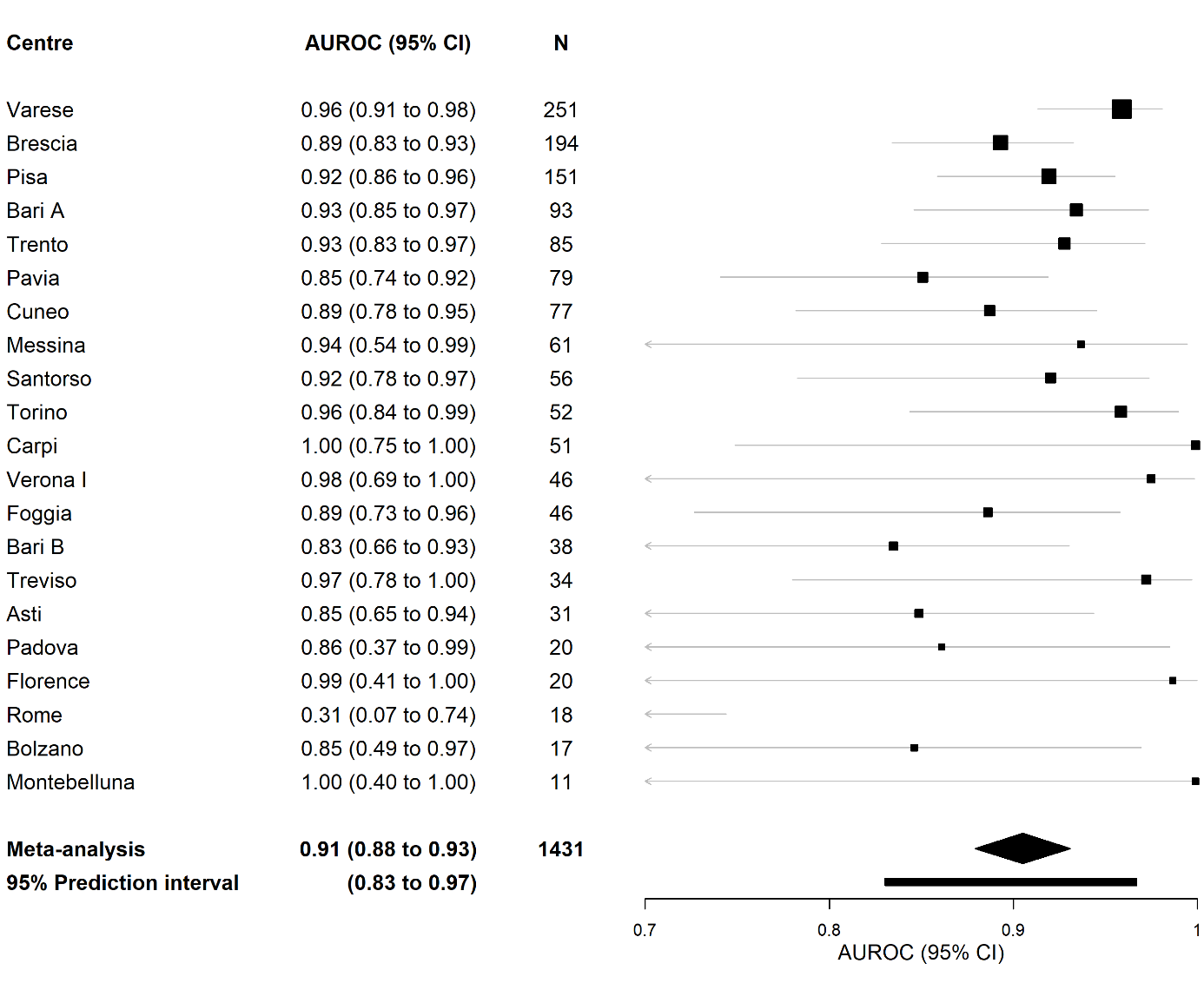

**Supplementary Figure S6**. Forest plot with center-specific area under the receiver operating characteristic curve (AUROC) of two-step strategy with CA125. For Rome, 2 tumors were borderline tumors and the rest were benign. The risk of malignancy estimated by the two-step strategy with CA125 was 1.83% and 1.65% for the borderline tumors. CI; Confidence Interval.

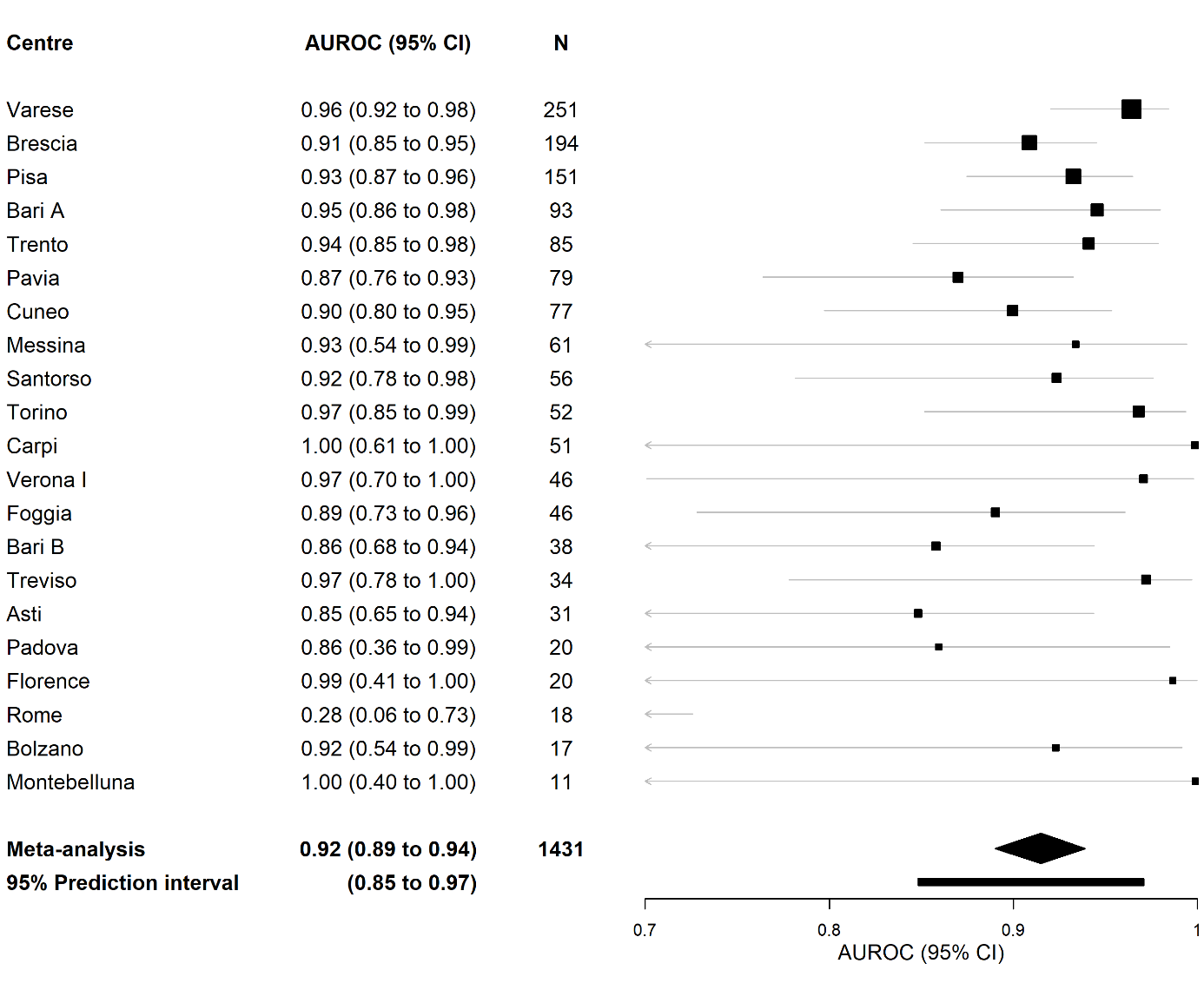

**Supplementary Figure S7**. Relation between the Risk of Malignancy Index (RMI) and the observed proportion of malignancy. Due to computational problems, we divided 13 centers with low sample size or low prevalence of malignancy into four groups: Santorso, Foggia, Treviso (group 1); Messina, Carpi, Montebelluna (group 2); Verona, Firenze, Padova, Roma (group 3); Bari B, Asti, Bolzano (group 4).The curve was obtained with meta-analysis of center-specific curves from eight centers and from the four groups. The dotted lines are the 95% confidence interval.

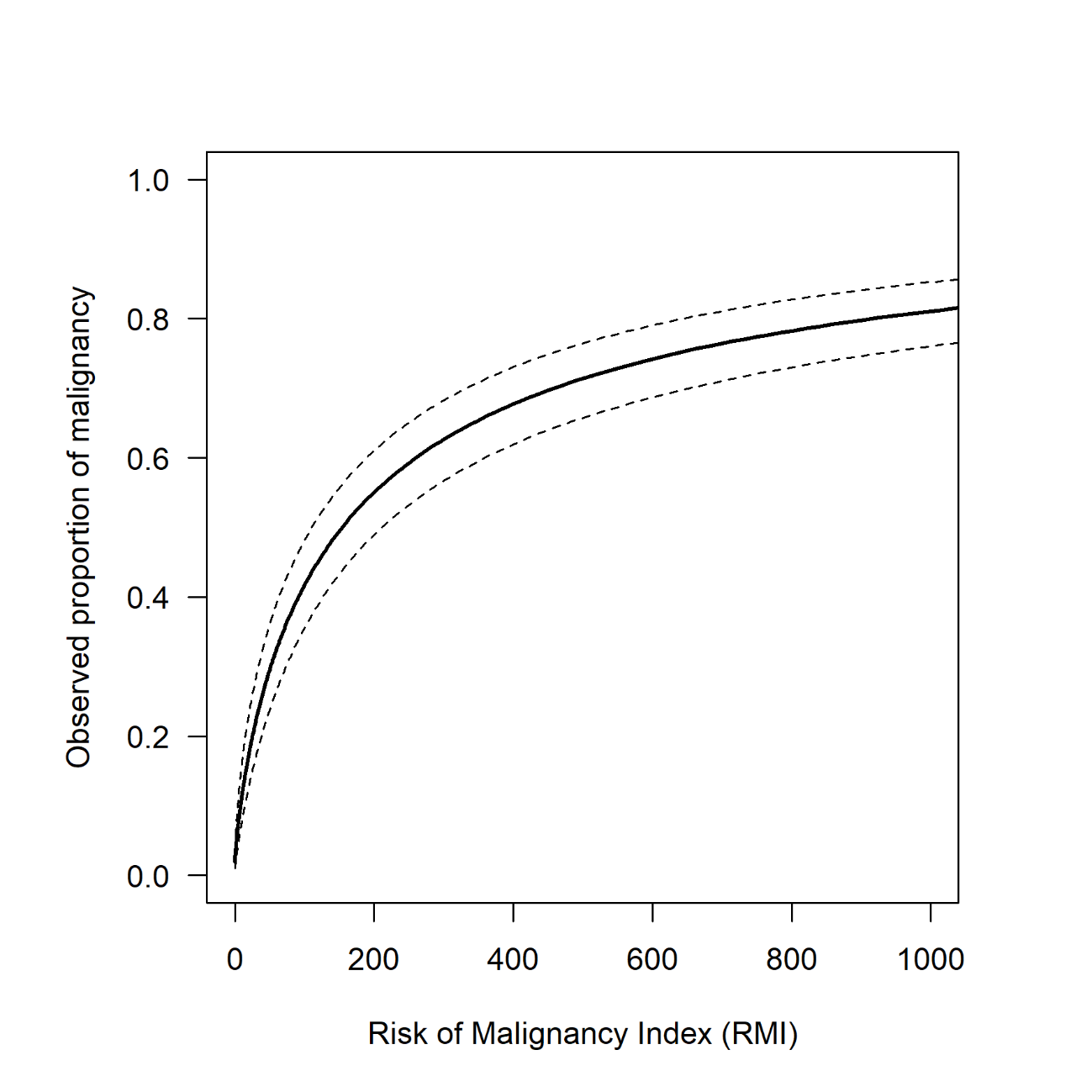

**Supplementary Figure S8**. Forest plot of O:E ratio for prespecified subgroups (pooled data). CI; Confidence interval, EFSUMB; European Federation of Societies for Ultrasound in Medicine and Biology.

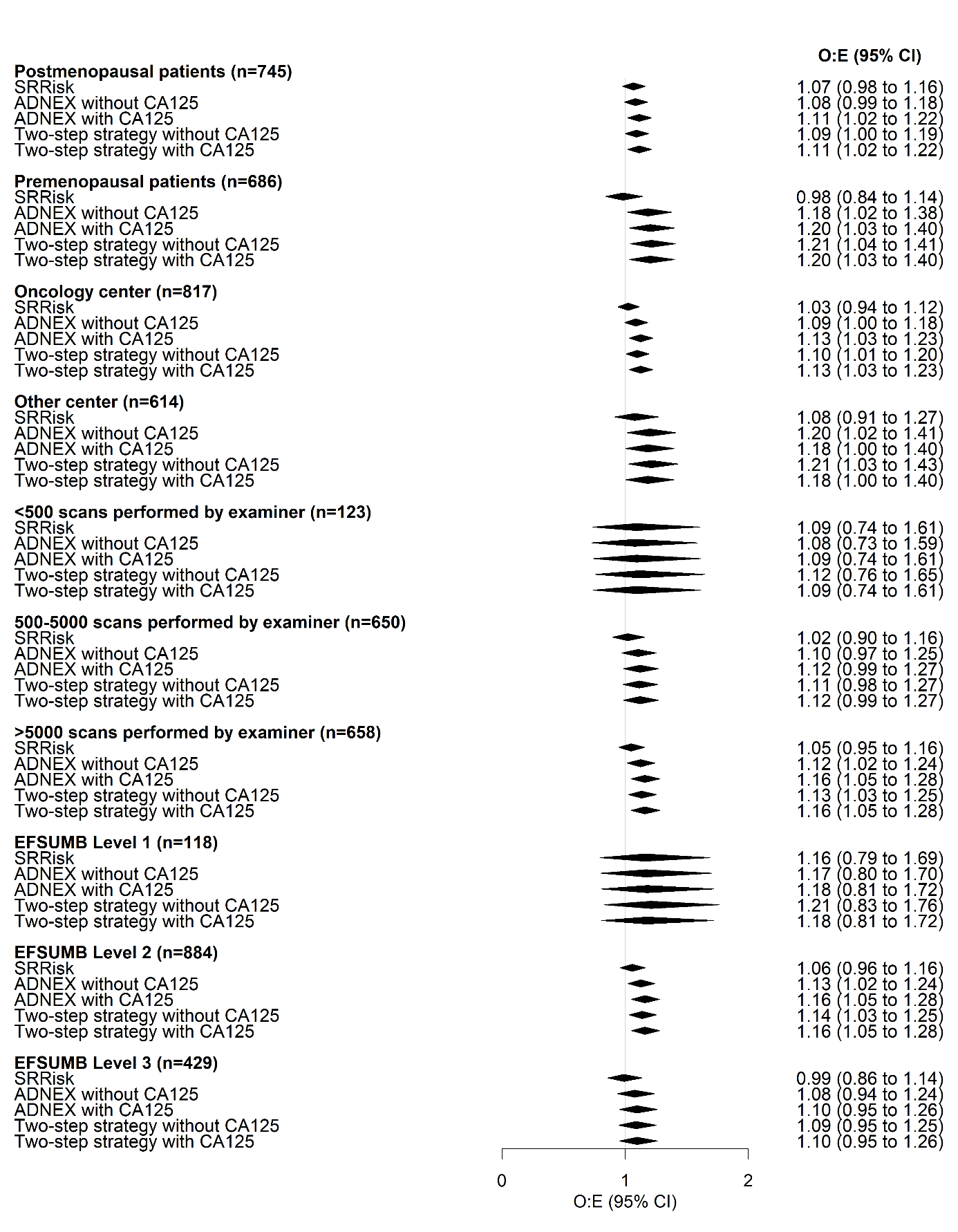

**Supplementary materials**

Appendix 1: Ethical Committee approval of each participating center

Cuneo n. 182-16, Bolzano n. 26, Brescia n. 2698, Asti n. 77-17, Bari n. 0070491, Messina n. 93/17, Verona n. 5186, Trento 1646/2017, Pisa n. 4335, Varese n. 11/2017, Torino n. 341, Carpi/Modena n. 0001285, Roma n. 319, Vicenza n. 62/18, Pavia n. 20180060503, Bari n. 176, Firenze n. 1725, Foggia n. 233, Abano Terme n. 2018 1519 E, Treviso n. 21466, Montebelluna n. 0129042

Appendix 2 : Estimated risks when Benign Descriptors apply

To assess the performance of the two-step strategy, risk predictions are needed. When no benign simple descriptor applies, the predictions of ADNEX are used. To assign a risk of malignancy to the Benign Descriptors we used the following data. Based on the results of IOTA phases 1-3 and of the immediately operated patients in the 2-year interim analysis of IOTA5, 13/1985 (0.65%) tumors to which a benign simple descriptor applied were malignant (i.e. 99.35% of these tumors were benign): 6/1985 (0.30%) were borderline tumors, 3/1985 (0.15%) were stage I primary ovarian malignancies, 1/1985 (0.05%) was a stage II-IV primary ovarian malignancy, and 3/1985 (0.15%) were secondary metastatic malignancies. These risks were used to assign a risk when the Benign Descriptors applied.

Appendix 3 : Sample size

The final IOTA6 protocol determined sample size as follows. The common rule of thumb for external validation studies is to have 100 observations in the smallest outcome groups. We expected that 10% of the patients selected for surgery at the primary centers would have a malignant tumor, which means that we would need 1000 patients.

We collected 1445 patients, including 1009 benign, 115 borderline, 106 stage I invasive, 154 stage II-IV invasive, 3 invasive with unknown FIGO stage and 58 metastatic tumors. Riley et al ^10^ have recently proposed a framework to calculate minimum sample size for external validation studies based on a specific targeted 95% CI width for calibration (Observed:Expected, O:E, and calibration slope), discrimination (C-statistic) and clinical utility (standardized Net Benefit, sNB). Table 7 shows the minimum sample size for different CI widths for different performance measures based on calculations described by Riley et al. In these calculations, a malignancy rate of 30% is used as observed in our dataset.

Calculations for calibration (O:E and calibration slope) suggest that at least 1401 (and 421 malignant tumors) are required to estimate an O:E with a 95% CI width of 0.16 assuming that the expected O:E is 1. 1476 participants (443 malignant cases) are required to estimate a calibration slope with a CI width of 0.20 assuming that the expected slope is 1. For the C-statistic, at least 1433 participants (including 430 malignant cases) are required to estimate an expected AUC of 0.80 with a 95% CI width of 0.05. For the net benefit, the minimum number of participants required varies from 217 (including 66 malignant outcomes) to estimate a net benefit at a threshold of 0.01 with a 95% CI width of 0.05 up to 1390 (and 417 malignant tumors) participants to estimate a net benefit at threshold 0.10 with a 95% CI width of 0.05. Therefore, our sample size of 1445 including 436 malignant outcomes is sufficient to estimate the O:E, sNB and C-statistic with reasonable precision. For the calibration slope the precision is lower.

**Table S2.1.** Sample size calculation

| **Performance measure** | **Assumed value** | **Targeted 95% CI width (SE)** | **Number of participants (events) required** |
| --- | --- | --- | --- |
| O:E | 1 | 0.16 (0.04) | 1401 (421) |
| C statistic | 0.95 | 0.05 (0.0127) | 374 (113) |
|  | 0.90 | 0.05 (0.0127) | 746 (224) |
|  | 0.85 | 0.05 (0.0127) | 1103 (331) |
|  | 0.80 | 0.05 (0.0127) | 1433 (430) |
| sNB | 0.97 (t=0.01) | 0.05 (0.0127) | 217 (66) |
|  | 0.92 (t=0.05) | 0.05 (0.0127) | 699 (210) |
|  | 0.86 (t=0.10) | 0.05 (0.0127) | 1390 (417) |
|  | 0.78 (t=0.20) | 0.10 (0.0256) | 781 (235) |
|  | 0.74 (t=0.30) | 0.10 (0.0256) | 954 (287) |
|  | 0.67 (t=0.4) | 0.10 (0.0256) | 1378 (414) |
|  | 0.61 (t=0.5) | 0.15 (0.038) | 869 (261) |
| Calibration slope | 1 | 0.20 (0.051) | 1476 (443) |

Appendix 4 : Multiple imputation

Despite blood samples for measurement of serum CA125 being mandatory, 394 patients (28%) had missing values for CA125. To deal with missing values for CA125, we performed multiple imputation. Imputations were created using the method of fully conditional specification with the mice package in R. We generated 100 imputations, leading to 100 completed datasets. To estimate the missing values for CA125, we used predictive mean matching regression using the outcome, variables that are probably related to either the level of CA125 itself, or to the unavailability of CA125. In the imputation procedure, we have imputed log(log(CA125+1)) to deal with the extreme skewness of CA125. The following variables were used: presumed endometrioma based on subjective assessment (yes/no), subjective assessment at inclusion (5 ordinal groups: certainly benign, probably benign, uncertain, probably malignant or probably borderline, certainly malignant or certainly borderline), age of the patient (in years), type of center (oncology center vs non oncology center), maximal diameter of the lesion (in mm, log transformed), proportion of solid tissue (calculated as the maximum diameter of the largest solid component in mm divided by the maximum diameter of the lesion in mm, with a linear and quadratic term), number of locules (1, 2-10, >10, other), number of papillations (ordinal variable, 0, 1, 2, 3, >3), presence of acoustic shadows (yes/no), presence of ascites (yes/no), presence of metastases (yes/no), bilaterality (yes/no), papillary height (mm), presence of papillary projections with blood flow (yes/no), color score of intratumoral flow (ordinal variable with four levels 1-4), echogenicity of cyst fluid (nominal variable with 6 levels: anechoic, homogeneous low-level, ground glass, hemorrhagic, mixed, no cyst fluid), multinomial reference standard, and irregular internal cyst wall (yes/no).

**Figure S3.1**. Convergence plot. The plot on the left shows the mean value of log(log(CA125+1)), and the plot on the right shows the standard deviation. Each imputation is represented by a colored line, and the x-axis represents the iterations.

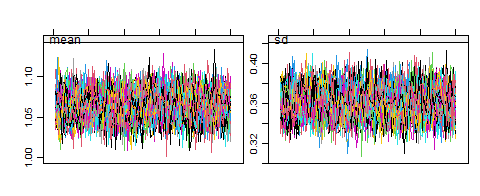

**Figure S3.2**. Density plot. The blue curve is the density of the observed values of log(log(CA125+1)), and the red curves are the densities for the imputed values of log(log(CA125+1)). There are 100 red lines, one for each imputation.

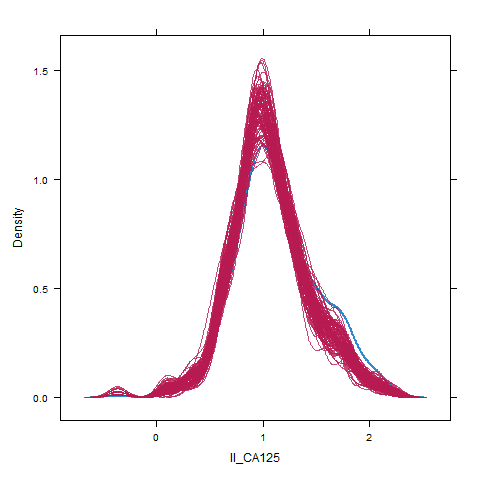

Appendix 5 : Details of the meta-analysis.

For the meta-analysis of AUROC, we obtained the logit(AUROC) and its standard error using the function auc.nonpara.mw function from the package auRoc. ^11^ 95% confidence intervals for logit(AUROC) were calculated, and then the point estimate and the confidence limits were back-transformed to the original scale. We obtained the overall AUC by combining center-specific logit(AUROC) and its standard error using random effects meta-analysis using the function valmeta from the package metamisc.^12^

To calculate O:E ratio, we calculated center-specific logit(O:E) and its standard error and combined them to obtain an overall O:E ratio using the function rma.uni from the package metafor. ^13^

We compute the multicenter calibration curves using a preliminary methodology based on a two-step meta-analysis. First, we train center specific flexible calibration models with LOESS. LOESS span is selected using fANCOVA r package that selects the best models using bias corrected AIC.^14^ Then for a fixed grid of values (e.g. 100 points from 0 to 1) we estimate the observed proportion with each of the center specific models and meta-analyze the results per grid value using random effects meta-analysis (with function metagen from meta package).^15^ This is a preliminary method that is currently under development and was presented at ISCB 2024 in Greece. You can find updated version of the methods in the OSF repository (<https://osf.io/erju9/?view_only=de8c921003a34931ac2ae4a71f85d66b>). For the meta-analysis of calibration curves, we divided 13 centers with low sample size or low prevalence of malignancy into four groups to avoid computational problems. We combined Santorso, Foggia, Treviso (group 1); Messina, Carpi, Montebelluna (group 2); Verona, Firenze, Padova, Roma (group 3); and Bari B, Asti, Bolzano (group 4).

Regarding clinical utility, we calculated net benefit (NB) for risk threshold between 1% and 50%. For each threshold, we calculated center-specific sensitivity, specificity and prevalence and combined them using Bayesian trivariate random effects meta-analysis to obtain an overall net benefit at each specific threshold using the package rjags.^16,17^

For meta-analysis of sensitivity, specificity, negative predictive value (NPV), and positive predictive value (PPV), we used a generalized linear mixed model (GLMM) with a logit link using the function metaprop from the package meta. ^15,18^ When a center had zero true positive (TP) and false positive (FP) at a specific threshold, it was excluded from the meta-analysis of PPV at that specific threshold. When a center had zero true negative (TN) and false negative (FN) at a specific threshold, it was excluded from the meta-analysis of NPV for that threshold.
